# Raised cardiovascular disease mortality after central nervous system tumour diagnosis: analysis of 171 926 patients from UK and USA

**DOI:** 10.1101/2021.03.29.21254512

**Authors:** Kai Jin, Paul M Brennan, Michael TC Poon, Cathie LM Sudlow, Jonine D Figueroa

## Abstract

**Background:** Patients with central nervous system (CNS) tumours have significant morbidity and mortality. Some studies showed CNS tumours patients may be at risk for cardiovascular disease (CVD) mortality. The magnitude of CVD risk among CNS tumours patients has not been comprehensively assessed. If CVD mortality is elevated in this population, there may be a potential for risk reduction to improve outcomes. We examined CVD mortality risk in patients with malignant and non-malignant CNS tumours.

**Methods:** We conducted analyses using UK (Wales)-based Secure Anonymised Information Linkage (SAIL) for 8,743 CNS tumour patients diagnosed in 2000-2015 (54.9% of whom died), and US-based National Cancer Institute’s Surveillance, Epidemiology, and End Results (SEER) for 163,183 patients in 2005-2015 (39.6% of whom died). We calculated age-, sex-, and calendar-year-adjusted standardised mortality ratios (SMRs) for CVD death in CNS tumour patients compared to Welsh and US residents. We used multivariable cause-specific Cox regression models to examine factors associated with CVD mortality among CNS tumour patients. We stratified all analyses by malignancy and main histological types.

**Results:** CVD was the second commonest cause of death for CNS tumour patients in SAIL (UK) and SEER (US) (9.5% & 11.7%, respectively). Patients with CNS tumours had higher CVD mortality than the general population (SAIL SMR=2.64, 95% CI=2.39-2.90, SEER SMR=1.38, 95% CI=1.35-1.42). Malignant CNS tumour patients had over 2-fold higher CVD mortality risk in both US and UK cohorts. SMRs for non-malignant tumours were almost 2-fold higher in SAIL than in SEER (SAIL SMR=2.73, 95% CI=2.46-3.02, SEER 1.30, 95% CI=1.26-1.33). The greatest magnitude of excess CVD mortality risk, particularly from cerebrovascular disease, was substantially greater in patients diagnosed at age younger than 50 years and within the first year after their cancer diagnosis (SAIL SMR=2.98, 95% CI=2.39-3.66; SEER SMR=2.14 95% CI=2.03-2.25). Age, sex, race/ethnicity in USA, deprivation in UK and no surgery were associated with CVD mortality.

**Discussion:** CVD mortality is high among patients diagnosed with both malignant and non-malignant CNS tumours compared to the general population. Targeted interventions and risk stratification tools might improve survival.

## Introduction

Central nervous system (CNS) tumours, both malignant and non-malignant, are associated with significant morbidity and mortality^1^. The incidence of CNS tumours is increasing globally^2^. Therapy options and patient survival for glioblastoma, the most common malignant CNS tumour, have changed little in almost 20 years, with median survival less than 15 months^3 4 5^. Meningioma, the most common non-malignant CNS tumour has 5-year survival of 70%^3^. There are substantial disabling effects associated with CNS tumours even if non-malignant, with a significant burden of disease for patients and their families, healthcare systems and societies worldwide^1^.

Accumulating evidence suggests cardiovascular disease (CVD) is the second leading cause of death in cancer patients. Up to 20% of cancer patients die from CVD, with risk varying by cancer type^6^. The potential causes of increased CVD risk in these patients include shared biologic mechanisms, modifiable risk factors, cancer treatment and access to healthcare^7-9^. In CNS tumours, CVD mortality is reportedly highest amongst younger (<40 years) cancer patients^8,10^. However, CVD mortality risk has not been systematically assessed in patients with CNS tumours. Previous studies were limited to small numbers of younger patients with benign CNS tumours^11-14^, or reported aggregated data for CNS tumours without distinguishing tumour subtypes^8,10,15^, which may have different effects on CVD mortality.

There has been no significant progress in improving survival for patients with CNS tumours in recent years with standard cancer therapy strategies^5^. Effective primary and secondary prevention of CVD including lifestyle changes and risk factor control have contributed to substantial reduction of CVD mortality worldwide in past three decades^16^, but may be underutilised among those with CNS tumours. If CVD mortality is elevated in patients with CNS tumours, prevention and treatment of CVD among those at high risk may improve outcomes.

We aimed to determine whether CVD mortality is raised among patients with different types of CNS tumours compared to the general population, and to assess the factors associated with any increased CVD mortality using population-based cancer registry data from UK and US.

## Methods

### Overall design

We conducted a retrospective, observational cohort study according to the Strengthening the Reporting of Observational Studies in Epidemiology (STROBE) guidelines^17^. First, we calculated standardised mortality ratios (SMRs) to assess the relative risk of CVD death for patients with CNS tumours compared to the general population, using data from UK (Wales)-based Secure Anonymised Information Linkage (SAIL) Databank^18^ and from US-based National Cancer Institute’s Surveillance, Epidemiology, and End Results (SEER) programme. Second, we examined the factors associated with CVD mortality in CNS tumour patients using the SAIL and SEER datasets. We provide more methodological detail in the supplemental file.

### Data sources

Both SAIL and SEER have been previously described^2,6^. Summary characteristics of the SAIL and SEER database are described in Online Table 1. Briefly, the SAIL Databank is a data platform holding de-identified and linkable datasets, including the Welsh Cancer Intelligence and Surveillance Unit (WCISU) data, for the population of Wales (approximately three million people), United Kingdom (UK). SEER is a registry of population-based incident tumour registries from geographically distinct regions in the US, covering 28% of the US population, including incidence, survival, and surgical treatment data^19^.

**Table 1.** Cohort characteristics in patients with central nervous system tumours.

| **SAIL 2000-2015** | **Overall CNS tumours** | **Non-malignant CNS tumours** | **Malignant CNS tumours** | **P-value** |
| --- | --- | --- | --- | --- |
|  | **N=8743** | **N=5119** | **N=3624** |  |
| **Sex** |  |  |  |  |
| **Female** | 4736(54.2%) | 3190(62.3%) | 1546(42.7%) |  |
| **Male** | 4007(45.8%) | 1929(37.7%) | 2078(57.3%) |  |
| **Age at diagnosis (years, mean/SD)** | 61.3(17.7) | 60.6(18.6)) | 62.2(16.4) | <0.001 |
| **Age at diagnosis** |  |  |  | <0.001 |
| **18-49 years** | 2207(25.2%) | 1466(28.6%) | 741(20.4%) |  |
| **Over 50 years** | 6536(74.8%) | 3653(71.4%) | 2883(79.6%) |  |
| **Area deprivation WIMD** |  |  |  | <0.006 |
| **1 Most deprived** | 1672(19.3%) | 1003(19.8%) | 669(18.7%) |  |
| **2** | 1688(19.5%) | 1002(19.8%) | 686(19.2%) |  |
| **3** | 1821(21.1%) | 1113(22.0%) | 708(19.8%) |  |
| **4** | 1787(20.7%) | 1000(19.7%) | 787(22.0%) |  |
| **5: Least deprived** | 1677(19.4%) | 950(18.7%) | 727(20.3%) |  |
| **Histologic subtypes** |  |  |  | <0.001 |
| **Glioma** | 3156(36.1%) | 83(1.6%) | 3073(84.8%) |  |
| **Meningioma** | 2128(24.3%) | 2036(39.8%) | 92(2.5%) |  |
| **Other** | 3459(39.6%) | 3000(58.6%) | 459(12.7%) |  |
| **Surgery** |  |  |  | 0.0998 |
| **Having surgery** | 5597(64.0%) | 3240(63.3%) | 2357(65.0%) |  |
| **No surgery or no information** | 3146(36.0%) | 1879(36.7%) | 1267(35.0%) |  |
| **Follow up time (months)\*** | 18.0[4.0; 68.4] | 42.6[11.0;92.6] | 6.0[2.0;18.0] | <0.001 |
| **Follow up period** |  |  |  | <0.001 |
| **< 1 year** | 3691(42.2%) | 1322(25.8%) | 2369(65.4%) |  |
| **1-4 years** | 2582(29.5%) | 1702(33.2%) | 880(24.3%) |  |
| **5-9 years** | 1554(17.8%) | 1318(25.7%) | 236(6.5%) |  |
| **≥10 years** | 916(10.5%) | 777(15.2%) | 139(3.8%) |  |
|  | **Total death** | **Total death** | **Total death** |  |
| **Main cause of death** | **N=4804** | **N=1738** | **N=3066** | <0.001 |
| **CNS death** | 3325(69.2%) | 549(31.6%) | 2776(90.5%) |  |
| **CVD death** | 455(9.5%) | 398(22.9%) | 57(1.9%) |  |
|  | **Total CVD death** | **Total CVD death** | **Total CVD death** |  |
| **Main types of CVD death** | **N=455** | **N=398** | **N=57** | <0.001 |
| **Heart disease** | 240(52.7%) | 208(52.3%) | 32(56.1%) |  |
| **Cerebrovascular disease** | 190(41.8%) | 167(42.0%) | 23(40.3%) |  |

| **SEER 2005-2015** | **Overall CNS tumours** | **Non-malignant CNS tumours** | **Malignant CNS tumours** | **P-value** |
| --- | --- | --- | --- | --- |
|  | **N=163183** | **N=110076** | **N=53107** |  |
| **Sex** |  |  |  | <0.001 |
| **Female** | 97944 (60.0%) | 74720 (67.9%) | 23224 (43.7%) |  |
| **Male** | 65239 (40.0%) | 35356 (32.1%) | 29883 (56.3%) |  |
| **Age at diagnosis (years, mean/SD)** | 60.7 (16.9) | 61.7 (16.6) | 58.8 (17.3) | <0.001 |
| **Age at diagnosis** |  |  |  | <0.001 |
| **18-49 years** | 40812 (25.0%) | 25943 (23.6%) | 14869 (28.0%) |  |
| **Over 50 years** | 122371 (75.0%) | 84133 (76.4%) | 38238 (72.0%) |  |
| **Race/Ethnicity** |  |  |  | <0.001 |
| **Non-Hispanic White** | 116509 (71.4%) | 76008 (69.1%) | 40501 (76.3%) |  |
| **Hispanic (All Races)** | 18522 (11.4%) | 12380 (11.2%) | 6142 (11.6%) |  |
| **Non-Hispanic Black** | 14421 (8.84%) | 11163 (10.1%) | 3258 (6.13%) |  |
| **Non-Hispanic Asian or Pacific Islander** | 11364 (6.96%) | 8599 (7.81%) | 2765 (5.21%) |  |
| **Non-Hispanic American Indian/Alaska Native** | 955 (0.59%) | 703 (0.64%) | 252 (0.47%) |  |
| **Non-Hispanic Unknown Race** | 1412 (0.87%) | 1223 (1.11%) | 189 (0.36%) |  |
| **Marital status** |  |  |  | <0.001 |
| **Married/Partner** | 88560 (54.3%) | 57539 (52.3%) | 31021 (58.4%) |  |
| **Single/Separated/Divorced** | 64138 (39.3%) | 44380 (40.3%) | 19758 (37.2%) |  |
| **Unknown** | 10485 (6.43%) | 8157 (7.41%) | 2328 (4.38%) |  |
| **Histologic subtypes** |  |  |  | <0.001 |
| **Glioma** | 50534 (31.0%) | 1705 (1.6%) | 48829 (91.9%) |  |
| **Meningioma** | 81431 (49.9%) | 80535 (73.2%) | 896 (1.7 %) | <0.001 |
| **Other** | 32128(19.11%) | 27836(22.3%) | 3382(6.4%) |  |
| **Surgery** |  |  |  | <0.001 |
| **Having surgery** | 82681 (50.7%) | 45641 (41.5%) | 37040 (69.7%) |  |
| **No surgery or no information** | 80502 (49.3%) | 64435 (58.5%) | 16067 (30.3%) |  |
| **Follow up time (months)\*** | 38.0 [14.0;78.0] | 54.0 [25.0;90.0] | 13.0 [4.00;33.0] | <0.001 |
| **Follow up period** |  |  |  | <0.001 |
| **< 1 year** | 7224 (32.8%) | 3289 (21.5%) | 3935 (58.4%) |  |
| **1-4 years** | 2469 (11.2%) | 1220 (7.97%) | 1249 (18.5%) |  |
| **5-9 years** | 903 (4.09%) | 778 (5.08%) | 125 (1.85%) |  |
| **≥10 years** | 11458 (52.0%) | 10026 (65.5%) | 1432 (21.2%) |  |
|  | **Total death** | **Total death** | **Total death** |  |
| **Main cause of death** | **N=64676** | **N=25042** | **N=39634** | <0.001 |
| **CNS death** | 33348 (51.6%) | 1081 (4.32%) | 32267 (81.4%) |  |
| **CVD death** | 7583 (11.7%) | 6525 (26.1%) | 1058 (2.67%) |  |
|  | **Total CVD death** | **Total CVD death** | **Total CVD death** |  |
| **Main types of CVD death** | **N=7583** | **N=6525** | **N=1058** | <0.001 |
| **Heart disease** | 5039 (66.4%) | 4354 (66.7%) | 685 (64.7%) |  |
| **Cerebrovascular disease** | 1990 (36.2%) | 1682 (25.8%) | 308 (29.1%) |  |

**\*Number presented in median/Interquartile range (IQR). CNS: Central nervous system; CVD: Cardiovascular disease includes the disease of heart, cerebrovascular disease, hypertension, atherosclerosis, aortic aneurysm/dissection, and other diseases of arteries, arterioles, or capillaries. WIMD: Welsh Index of Multiple Deprivation is the measurement of area deprivation in Wales, UK, ranking from 1(most deprived) to 5 (least deprived).**

### Study population

We identified patients diagnosed with CNS tumours in 2000-2015 from SAIL and SEER in 2005-2015. We classified CNS tumours using the following International Classification of Diseases 10^th^ revision (ICD-10) codes: C70-72, C75.1-75.3, D18.0, D32-D33, D35.2-D35.4, D42-D43, D44.3-D44.5^2^. We also used the International Classification of Diseases for Oncology third edition (ICD-O-3) codes to group tumours into malignancy type (malignant or non-malignant type) and three major histologic subgroups (meningiomas, gliomas, and all others), following the definitions from the Central Brain Tumour Registry of the United States (CBTRUS) (Online Appendix 1)^3^. We restricted analysis to adults aged ≥18 years at date of diagnosis.

### Outcome variables

We ascertained deaths by linking to mortality data and categorised causes of death using ICD-9 and ICD-10 codes for the main underlying cause: CNS tumours, cardiovascular disease (CVD) and other causes (Online Appendix 2 & 3). The primary outcome of interest was death from CVD, including diseases of the heart, cerebrovascular disease, atherosclerosis, and aortic aneurysm or dissection^6^.

### Statistical analysis

We compared CVD mortality among patients with CNS tumours to the general population using standardised mortality ratios (SMRs), calculated as the ratio of the observed to expected numbers of deaths. We calculated these for the time period 2000-2015 for SAIL UK and 2005-2015 for SEER US, accounting for the fact that non-malignant CNS tumours were only registered in SEER from 2004 onwards^20^. We adjusted SMRs by age, sex, and calendar year to the Welsh and US general populations respectively during the study period. For both datasets, we categorised age groups as 18-49 years (due to low frequency of cases in younger people) and into 10-year age groups for ages ≥50 years. We estimated SMRs for all CVD deaths and for two major CVD subgroups (deaths due to heart disease and those due to cerebrovascular disease) as a function of calendar year, age at cancer diagnosis, and follow-up time after cancer diagnosis, respectively.

To determine factors associated with elevated CVD mortality in CNS tumour patients, we used cause-specific multivariable Cox proportional hazards regression models to calculate hazard ratios (HRs) and 95% confidence intervals (CIs) for CVD deaths as the primary outcome among different CNS tumour patient subgroups. The variables included in the models were age at diagnosis, malignancy (malignant and non-malignant type), histological types (gliomas, meningioma, and other histological types), age at diagnosis, sex, race/ethnicity status (SEER only), marital status (SEER only), area deprivation measured by Welsh Index of Multiple Deprivation (WIMD) (SAIL only) and surgery status (surgery versus no surgery or no information). We computed survival time from the date of CNS cancer diagnosis until date of death or last contact (December 31, 2015). We calculated cumulative incidence function curves to describe the incidence of deaths from CVD, and CNS death over time. We also conducted analyses stratifying CNS tumour patients by malignant status and major histological subtypes. To reduce the chance of reverse causality, all analyses were landmark analyses, with follow-up commencing 2 months after cancer diagnoses^15^, thereby excluding patients with an event of death from cancer or CVD within 2 months. To test the robustness of our results, we performed sensitivity analyses in those diagnosed from 2005 onwards when incidence and diagnosis of CNS tumours (overall and histologically confirmed) in SAIL stabilised^2^.

We defined statistical significance by a two-tailed p-value <0.05. We prepared and analysed data using R (Version 4, https://www.R-project.org/) for SAIL UK and SEER*Stat 8.3.8 for SEER. We conducted analyses of SAIL data within SAIL’s remotely accessible trusted research environment, having obtained approval for use of the data (Project No. 0918). No additional informed consent was required as there was no individual patient involvement.

## Results

### Cohorts characteristics

Table 1 shows characteristics overall and by main causes of death in SAIL and SEER cancer registries. There were 8,743 patients with CNS tumours diagnosed between 2000 and 2015 in SAIL with 30,134 person-years at risk. Nearly 60% (n=5,119) of patients had non-malignant tumours, which were more frequent in women. More men than women had a malignant tumour. The most common histological subtypes were gliomas (36.1%), which are mostly malignant, and meningiomas (24.3%), the majority of which are non-malignant. Among 4,804 (54.9%) deaths recorded in SAIL, CNS tumour was the leading cause of death (69.2%) followed by CVD (9.5%). The primary cause of CVD death was heart disease (52.7%), followed by cerebrovascular disease (41.8%).

In SEER, there were 163,183 patients with CNS tumours diagnosed between 2005 and 2015 with 684,437 person-years at risk (Table 1). Over 67% (n=110,076) of patients had non-malignant tumours, which were more frequent in women. The most common histological subtypes were meningioma (49.9%) and glioma (31.0%). Among 64,676 deaths, CNS tumour was the leading cause of death (51.6%) followed by CVD (11.7%). The primary cause of CVD death was heart disease (66.4%), followed by cerebrovascular disease (36.2%).

### Cardiovascular mortality risk

Figure 1 presents CVD mortality risk in SAIL and SEER. Overall, patients with CNS tumours had higher CVD mortality compared with the general population (SAIL SMR=2.64, 95% CI=2.39-2.90, SEER SMR=1.38, 95% CI=1.35-1.42). Malignant CNS tumour patients had more than twice the CVD mortality risk of the general population in both the UK and the US (Figure 1). SMR for non-malignant tumours was almost twice as high in SAIL as in SEER (SAIL SMR=2.73, 95% CI=2.46-3.02, SEER 1.30, 95% CI=1.26-1.33, Figure 1). SMRs for cerebrovascular mortality were higher than for heart disease mortality, with relative risk considerably higher in SAIL than in SEER (Figure 1). Analysis of mortality trends by calendar year showed elevated CVD mortality risks across the entire period of study, albeit with some variation, in both SAIL and SEER (Online Table 2).

**Table 2.** Cox proportional hazards regression multivariable HRs and 95% CIs for association between patient characteristics and CVD mortality.

| **SAIL UK 2000-2015** | **Meningioma** | **Glioma** |
| --- | --- | --- |
| **Characteristics** | **HR(95% CI)** | **HR(95% CI)** |
| **Gender** |  |  |
| Female | 1 |  |
| Male | 1.16(0.85-1.57) | 2.11(1.03-4.31)* |
| **Age** | 1.11(1.09-1.12) † | 1.08(1.05-1.11) † |
| **Area deprivation** |  |  |
| WIMD 5(Least deprived) | 1 |  |
| WIMD 4 | 1.10(0.70-1.71) | 0.74(0.28-1.94) |
| WIMD 3 | 1.32(0.84-2.05) | 0.81(0.30-2.19) |
| WIMD 2 | 1.73(1.12-2.66)* | 0.63(0.21-1.9) |
| WIMD 1 (Most deprived) | 2.00(1.30-3.06) ‡ | 1.37(0.54-3.48) |
| **Calendar year** | 0.98(0.86-1.11) | 0.91(0.83-1.00) |
| **Treatment** |  |  |
| Having surgery | 1 | 1 |
| Not having surgery or no information | 1.33(0.99-1.79) | 2.22(1.03-4.76)* |
| **SEER 2005-2015** | **Meningioma** | **Glioma** |
| **Characteristics** | **HR (95% CI)** | **HR(95% CI)** |
| **Gender** |  |  |
| Female | 1.00 | 1.00 |
| Male | 1.64(1.55-.74) † | 1.36(1.18-1.56) † |
| **Age** | 1.10(1.10-.10) † | 1.07(1.07-1.08) † |
| **Ethnicity** |  |  |
| Non-Hispanic White | 1.00 | 1.00 |
| Hispanic(All races) | 0.91(0.83-1.01) | 1.10(0.87-1.39) |
| Non-Hispanic Black | 1.48(1.37-1.60) † | 2.01(1.61-2.53) † |
| Non-Hispanic Asian or Pacific Islander | 0.83(0.75-0.94) ‡ | 0.98(0.71-1.35) |
| Non-Hispanic American Indian/Alaska Native | 0.84(0.54-1.30) | 1.72(0.77-3.86) |
| Non-Hispanic Unknow race | 0.20(0.08-0.48) † | 0.43(0.06-3.04) |
| **Marital status** |  |  |
| Married/Having Partner | 1.00 | 1.00 |
| Single/Separate/Divorced | 1.38(1.30-1.47) † | 1.43(1.23-.65) † |
| Unknown | 1.26(1.14-1.40) † | 1.21(0.87-1.70) |
| **Year of diagnosis** | 0.99(0.98-1.00)* | 0.96(0.94-0.99) ‡ |
| **Treatment** |  |  |
| Having surgery | 1.00 | 1.00 |
| Not having surgery or no information | 1.33(1.25-1.43) † | 1.56(1.35-1.56) † |
| **Meningioma: non-malignant meningioma, Glioma: malignant glioma; HR: Hazard ratio; CI: Confidence interval; CVD: Cardiovascular disease. WIMD: Welsh Index Multiple Deprivation \*<0.05, †<0.001, ‡ <0.01;** |  |  |

**Figure 1.**
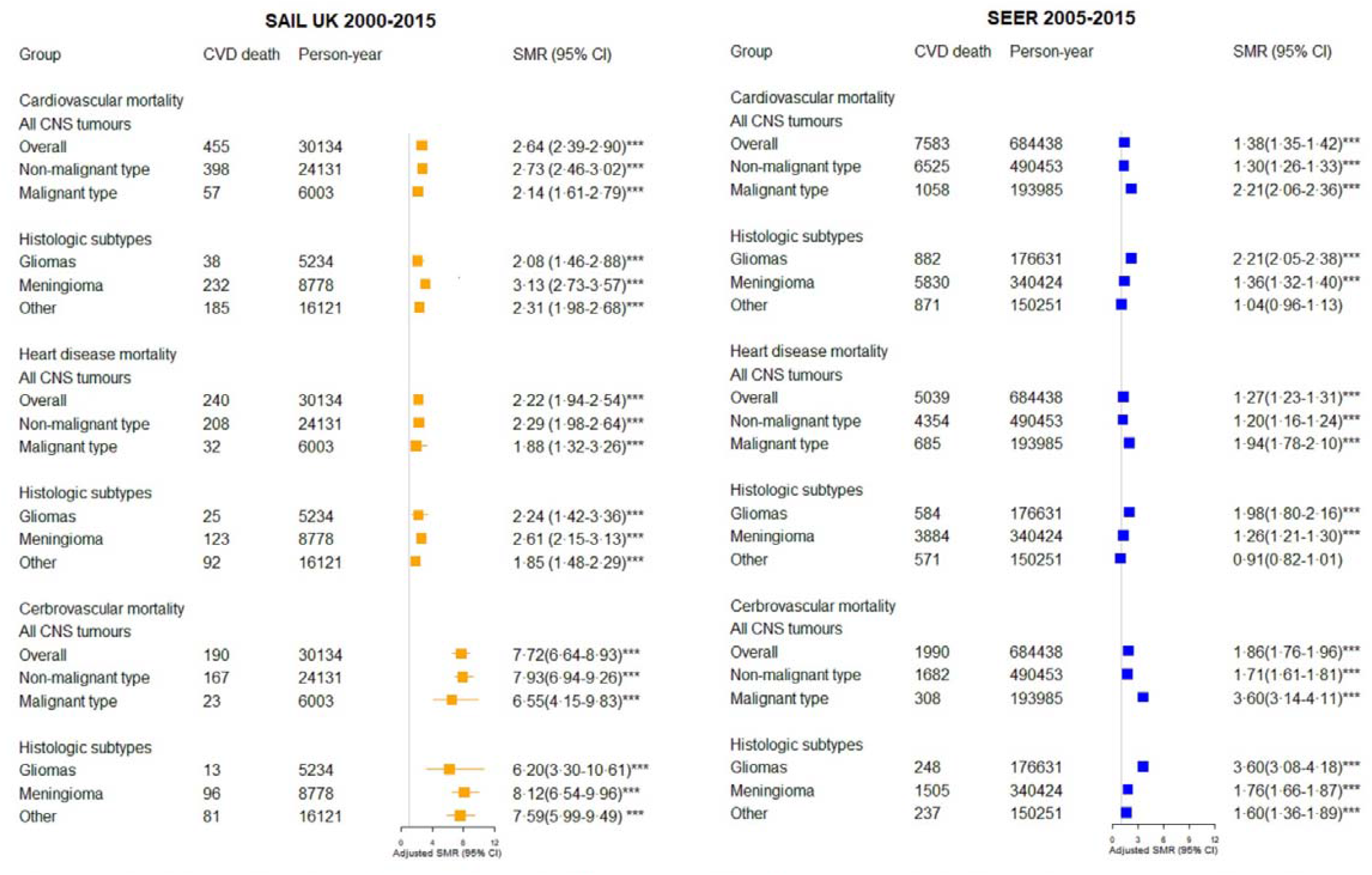
Cardiovascular mortality in CNS tumours and stratified by subgroups. Standardised mortality ratios (SMR) for cardiovascular disease (CVD) adjusted by age, sex and calendar-year in CNS tumours, and by malignancy and main histological subtypes. SMRs for subtypes within cause of CVD (disease of heart and cerebrovascular disease) were presented. A SMR above 1 represents a higher relative risk of death for a type of cardiovascular cause compared witli the general population. *** indicates p<0.0001

### Cardiovascular mortality risk by age at cancer diagnosis

For all CNS tumours combined, SMRs for CVD mortality were elevated for all age groups in SAIL and SEER, particularly for patients diagnosed below the age of 50 years (SAIL SMR=96.09, 95%CI=48.89-164.55, SEER SMR 2.03, 95%CI=1.79-2.30; Figure 2 & Online Table 3). These patterns of higher relative CVD mortality risk for those with younger age at diagnosis were consistent across CNS tumour subtypes. SMRs for cerebrovascular disease were substantially elevated in SAIL (SMR 321.97, 95%CI= 138.34-622.23), less so for SEER (SMR 4.82, 95%CI=3.84-5.99) (Online Table 3).

**Figure 2.**
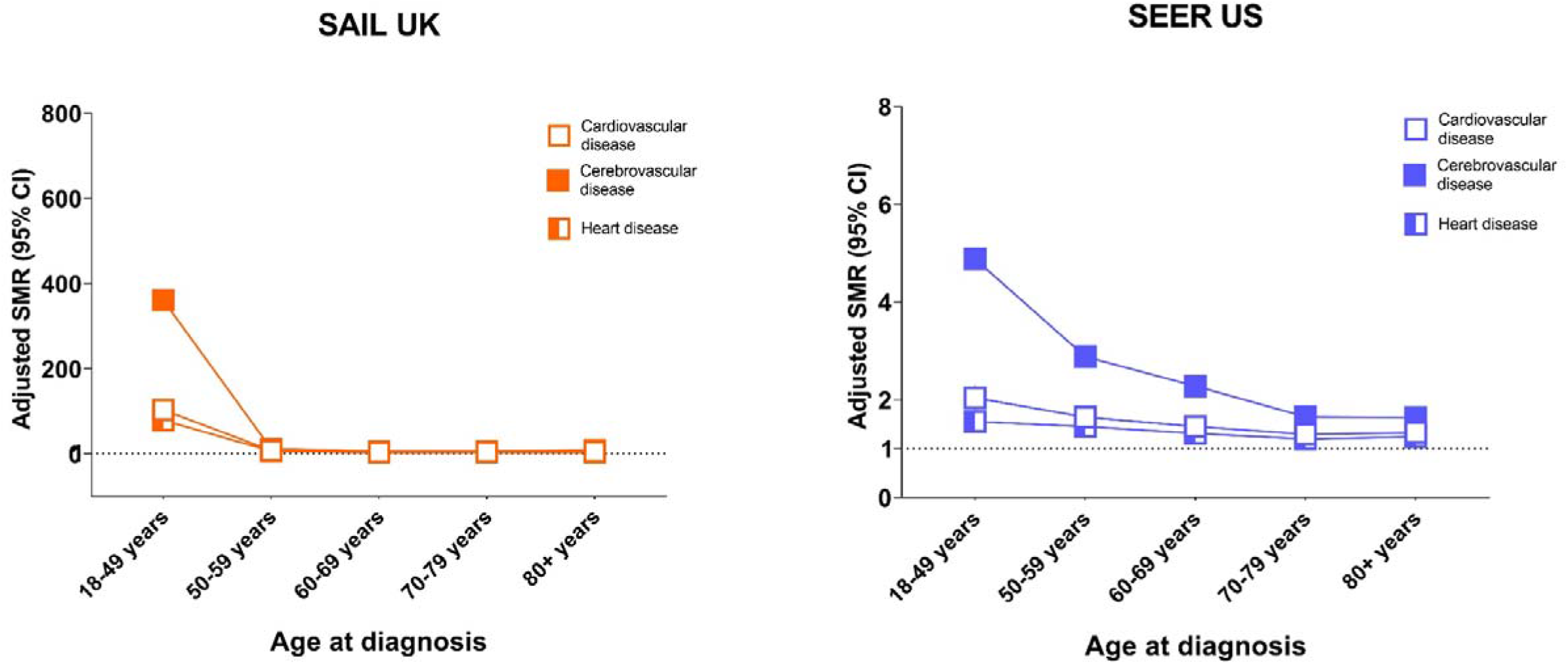
Cardiovascular mortality in CNS tumours by age at diagnosis and stratified by subtypes. Cardiovascular mortality by age at diagnosis in central nervous system (CNS) tumour patients. Standardised mortality ratios (SMR) and 95% confidence interval (CI) for CVD were adjusted by sex and calendar year. Relative risk of main causes of death within CVD (heart disease and cerebrovascular disease) were presented. A SMR above 1 represents a higher relative risk of death for a type of cardiovascular cause compared with the general population.

### Cardiovascular mortality risk during follow-up

The highest SMRs for CVD mortality were observed in the first year after cancer diagnosis (SAIL SMR=2.98, 2.39-3.66, SEER SMR=2.14, 2.03-2.25). CVD mortality declined from one year post diagnosis, but remained elevated for up to 10 years (Figure 3, Online Table 4). These patterns of higher CVD mortality risk during follow-up period after cancer diagnosis were consistent across CNS tumour subtypes.

**Figure 3.**
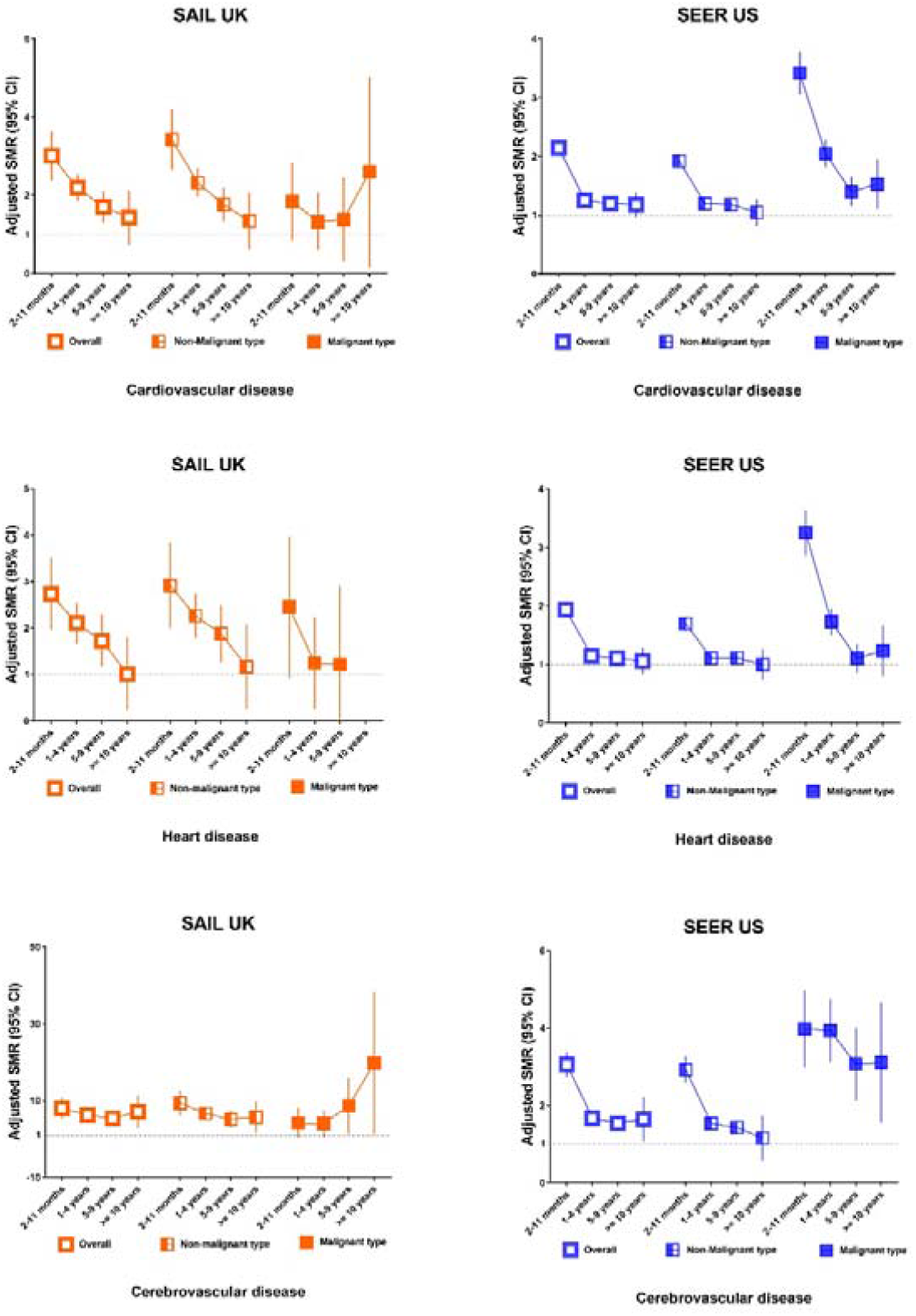
Cardiovascular mortality in CNS tumours by follow up year and stratified by subgroups. Cardiovascular mortality by follow up after cancer diagnosis. Standardized mortality ratios(SMRs) for cardiovascular disease adjusted by age, sex and calendar year in overall CNS tumours and by malignancy. SMRs for heart disease and cerebrovascular disease were presented. A SMR above 1 represents a higher relative risk of death for a type of cardiovascular cause compared with the general population.

### Cumulative incidence of death

The cumulative incidences of death from CVD and from CNS tumour by main histological group in SAIL and SEER are shown in Online Figure 1. The risk from CVD death was negligible in gliomas in both datasets but constituted the leading cause of death in meningioma in SEER.

### Factors associated with CVD mortality in SAIL and SEER

Multivariable Cox proportional hazards models to examine associations of demographic and clinical factors with CVD mortality in non-malignant meningioma and malignant gliomas are show in Table 2. For meningiomas, strong predictors of CVD mortality were older age and deprivation (SAIL), non-Hispanic Black ethnicity (SEER) and not being married or having a partner (SEER). Being male and not having surgery were associated with higher CVD mortality in both cohorts but non-significantly (P>0.05) in SAIL (Table 2). Notably, individuals from more deprived areas in SAIL had 73-100% higher risk of CVD death compared to those from the least deprived areas. For gliomas, being male, older age, and not having surgery significantly increased risk of CVD mortality in both cohorts. In the SEER cohort, being non-Hispanic Black or not being married or having a partner were also associated with higher CVD mortality. Not having surgery was associated with increase in risk of CVD death in SAIL (HR=2.22, 1.03-4.76) and SEER (HR=1.56, 1.35-1.56) respectively, compared with those having surgery (Table 2).

All-cause and CVD-cause mortality for overall CNS tumours and across malignancy subgroups are shown in Online Table 5. Overall, age, sex, race/ethnicity in USA, deprivation in the UK and not having surgery were associated with all-cause and CVD mortality in all CNS tumours combined. Having a non-malignant (versus malignant) CNS tumour was strongly associated with increased risk of CVD mortality in SAIL. In contrast, having a non-malignant tumour in SEER was associated with reduced CVD mortality.

## Discussion

### Main Findings

Our study is the largest population-based study to characterise CVD mortality in CNS tumour patients. Among over 170,000 patients with malignant and non-malignant CNS tumours, CVD was the second commonest cause of death. There was a significantly increased risk of CVD mortality after CNS tumour diagnosis compared to the general population. CVD mortality, particularly from cerebrovascular disease, was substantially greater in patients diagnosed at a younger age (<50 years) and within the first year after their cancer diagnosis. Age, sex, race/ethnicity in USA, deprivation in UK and not having surgery were associated with CVD mortality in CNS tumour patients. Targeted interventions and risk stratification tools might improve survival.

Our inclusion of all CNS tumour subtypes extends previous studies of CVD mortality among patients of younger age at onset (<40 years)^8,11,14^. Systematic review of previous studies comparing CVD mortality in CNS tumours patients with the general population can be found in the Supplement file (Part 4). Increased CVD mortality in malignant CNS tumour patients may reflect cardiotoxicity from cancer treatment, their associated health conditions, or lower frequency of use of treatments for preventing CVD. Current treatment for malignant CNS tumours includes surgery, cranial radiotherapy, and chemotherapy^4^. Cancer treatment is associated with adverse cardiovascular complications including cardiomyopathy, cardiac arrhythmias and stroke^21^. Older age also contributes to increased risk of cardiotoxicity from cancer treatment^6^, and indeed, the mean age at cancer diagnosis was 60 years in our cohorts. Further research should establish whether preventive therapies could reduce this risk. Our study provides information to inform CVD risk stratification that could guide this work. Increased CVD mortality risk in non-malignant CNS tumour patients, who do not receive anti-cancer treatment, could relate to existing CVD or other comorbidities, age, or reduced use of or access to existing preventive therapies^22^, and requires further study.

We observed that CVD mortality risk declined over the decades studied. Similar trends have been reported in CVD mortality risk among patients from other non-CNS cancers^8,23^. This may reflect contemporary efforts to minimise exposure to cardiovascular adverse effects from chemotherapy in cancer patients, as well as improvements in CVD preventive treatments more broadly^24^.

Our finding that patients who are younger at cancer diagnosis, and in the first year after diagnosis, were at higher risk of CVD mortality is consistent with previous studies of other cancers^6,8,15^. In one study following over 200,000 young adult cancer patients for 25 years, risk of cardiac mortality was 1.4 times the general population with decreasing SMRs for cardiac mortality at increasing ages (SMR 4.2 at 15-19 years to SMR 1.2 at 35-39 years)^8^. This risk of death from CVD after cancer treatment remained high for at least 25 years. We found that CVD mortality risk remained elevated up to 10 years after diagnosis. The immediate negative effects of surgery (malignant and non-malignant tumours), and concomitant radiotherapy and chemotherapy (malignant tumours) may explain the highest risk in first-year post diagnosis.

Across all CNS tumours, cerebrovascular mortality risk was higher than heart disease. Greater cerebrovascular mortality risk was reported in previous studies of childhood and young adult onset CNS tumours^10,11,25^. A recent US study found malignant CNS tumours had the highest risk of stroke mortality(SMRs 2-5) compared to other cancer sites in the first five years following cancer diagnosis^15^. Cranial irritation from radiotherapy along with neurosurgery could contribute to the higher cerebrovascular risk in malignant CNS tumour patients^11,26^. In support of this, cancers such as breast cancer and Hodgkin lymphoma, radiotherapy to the thorax leads to high risk of ischaemia heart disease^8^. However, the higher stroke mortality may also be partly explained by direct effects of CNS tumours on cerebral vasculature, systemic tumour-related inflammatory and pro-coagulant factors, or – in some cases – diagnostic misclassification (e.g., focal epilepsy, ‘tumour attacks’, or intra-tumoural haemorrhage being labelled diagnostically as stroke).

Our results suggest that not having surgery is associated with increased all-cause mortality and CVD mortality. This may implicate a tumour-related factor in CVD risk but may alternatively indicate that more proactive follow up of patients after surgery is of benefit for earlier identification and treatment of CVD risk factors. Another explanation is ‘reverse causality’, that those who did not receive surgery had worse performance status or poor neurological function at the time surgery was considered, particularly in elderly patients^27^. The cohorts we examined lacked the granular clinical detail to enable further exploration of these potential explanations.

CVD mortality risk varied between the UK and US cohorts studied, especially for meningiomas. Meningioma patients in SAIL had much higher CVD mortality risk than those in SEER (SMRs 3.13 and 1.36 respectively). Direct comparison of these cohorts is difficult, because of geographic differences in incidence, access to specialised medical and surgical care for diagnosis and long-term management, and socio-demographics, all of which could lead to substantial differences in risk^8,28^. Highly specialised neuro-oncology care including surgical and radiation services for diagnosis and long-term management are important for improving survival outcome in CNS tumours^1^. Incidence rates of meningiomas in the US are higher than in other countries, including the UK^2,28^. This likely reflects increasing diagnosis of meningiomas as incidental findings, partly because of growing MRI availability, more marked in the US^28^. More meningiomas are diagnosed and at an earlier stage^29^. The rate of growth of meningioma is typically slow and most meningiomas found incidentally on brain imaging undertaken for unrelated symptoms remain asymptomatic^29^. A larger proportion of meningiomas detected in the US when they are small, not growing, and not requiring treatment, may at least in part explain the difference in excess CVD mortality between the UK and US. A better understanding of the factors contributing to CVD risk following a non-malignant CNS tumour diagnosis is needed to interrogate the difference further. Less well-resourced healthcare and higher prevalence of CVD risk factors can compound CVD mortality risk^30^, and may also contribute to the differences observed between the US and the UK.

### Implications

Our study showed CVD mortality is the largest single non-cancer cause of death among patients with malignant and non-malignant CNS tumours in two independent cohorts, contributing to significantly worse CNS cancer survival. Our findings have important implications. Over 25% of CNS tumour patients were diagnosed aged 18-49 years, among which over 60% had a non-malignant tumour. This is important because these patients have better survival prognosis overall and so may have most to benefit from early CVD prevention. Malignant CNS tumour patients have, on average, a much shorter-life expectancy, but there still may be advantages to CVD risk reduction, especially in the absence of other emerging therapeutic strategies. Early prevention of CVD should be a priority in cancer patients. In our study, advanced age, being male, lower SES and non-Hispanic Black ethnicity were strong predictors of CVD mortality in CNS tumours patients. These are well established CVD mortality risk factors and could inform screening to identify patients who may benefit from CVD preventive interventions. Active monitoring and treatment of CVD complications during and after cancer treatment could also help reduce CVD mortality.

### Study Strengths and Limitations

Our results provide the most comprehensive analysis of CVD mortality in CNS tumour patients, enhancing the generalisability of our findings. Using cancer registry data from two countries allowed us to examine the CVD mortality patterns across relatively rare CNS tumour subtypes using consistent methodology. Population-based cancer registry data has strengths of capturing and identifying risk in those who may be under-represented in clinical trials, from which treatment guidelines are often developed. Clinical trials in malignant tumours such as glioblastoma usually recruit patients with better performance status, often excluding older individuals with multi-comorbidities.

Our study had several limitations. Firstly, SMRs should be interpreted with caution since they compare the relative risk with the reference population, which may vary among groups and geographical location^15^. The precision of SMRs depends on person-years at risk, and if the incidence of a cancer is low (e.g. gliomas) and survival is limited, CIs may be wide. Second, we lacked detailed information on exposure to the most relevant treatment modalities, such as specific type of surgery, chemotherapy, and radiotherapy, limiting further exploration and interpretation of our results. We had no reliable information on cancer recurrence, which could be an important predictor of further cancer treatment. Third, we did not have information on CVD risk factors such as obesity and smoking. However, a recent large study examining CVD risk for multiple cancer sites showed adjustment for shared risk factors had little effect on CVD risk compared with the general population^25^. Fourth, the lack of individual level of sociodemographic information such as marital status and ethnicity in SAIL and deprivation in SEER made it difficult to directly compare CVD risk for these two cohorts. Another limitation is the use of death certificate information to classify cause of death, which may have limited accuracy. However, international studies have reported acceptable validity (83-98%) of case of death using such data^23,31^. Lastly, we did not have detailed data in SEER to classify all stroke outcomes as ischaemic or haemorrhagic.

## Conclusions

Our findings contribute to the understanding of CVD risk in CNS tumour patients. Despite advances in cancer treatment, mortality reductions over time from CNS tumours have been modest. Our results provide important evidence on CVD risk in CNS tumours by subtype that could inform risk stratification and prevention efforts in clinical practice, particularly in the first year and among those diagnosed at younger ages. Given the effectiveness of both primary and secondary prevention interventions for CVD, risk screening prior to, during and after cancer treatment should be emphasised and communicated within multidisciplinary teams including neuro-oncologists, general practitioners, and cardiovascular specialists.

## Data Availability

Data was from UK (Wales)-based Secure Anonymised Information Linkage (SAIL) Databank, and from US-based National Cancer Institutes Surveillance, Epidemiology, and End Results (SEER) programme.

## Acknowledgements

This study makes use of anonymized data held in the Secure Anonymised Information Linkage (SAIL) Databank. We would like to acknowledge all the data providers who make anonymized data available for research.

