## Supplemental file for "Raised cardiovascular disease mortality after central nervous system tumour diagnosis: analysis of 171 926 patients from UK and USA"

#### Table of Contents

Part 1: Expanded methods ………………………………………………………….Page 3

Part 2: Expanded materials…………………………………………………………… Page 6

Part 3: Online Tables and Figures ……………………………………………….....Page 8

Part 4: Systematic review of cardiovascular mortality in central nervous system tumours patients………………………………………………………………………. Page 22

### Part 1: Expanded methods

#### Overall design

This is a retrospective, observational cohort study according to the Strengthening the Reporting of Observational Studies in Epidemiology (STROBE) guidelines(8). Our analysis involved two main components. First, we calculated standardised mortality ratios (SMRs) for CVD in CNS tumours, using data from Secure Anonymised Information Linkage (SAIL) Databank (9) (https://saildatabank.com and data from the National Cancer Institute's Surveillance, Epidemiology, and End Results (SEER) program. SMRs provide the relative risk of CVD death for patients with CNS tumours as compared to the general population. Second, we examined the factors associated with CVD mortality in CNS tumours patients using SAIL and SEER datasets.

#### Data Source

##### SAIL UK

Patients with malignant and non-malignant CNS tumours, diagnosed between 2000 and 2015, were abstracted from SAIL. The SAIL Databank is a data platform holding de-identified and linkable datasets including Welsh Cancer Intelligence Surveillance Unit (WCISU), Welsh Mortality data, Welsh demographic data in Wales, United Kingdom (UK) with approximately three million people. The SAIL Databank is the representative of all population of Wales UK. WCISU is the national cancer registry for Wales, 100% representative of Welsh population, and includes data on sex, age at diagnosis, year of diagnosis, and treatment information(10). The causes of death were ascertained after linking to Welsh mortality data from Office for National Statistics (ONS) UK. Mortality statistics reported by ONS are based on information recorded when deaths are certified and registered in England and Wales. The death certificate used in England and Wales is compatible with WHO recommendation. Underlying cause of death is based on International Classification Code (ICD) rules, and is made from the conditions reported by the certifier, as recorded on the death certificate (User guide to mortality statistics, Office National Statistics 2019). The coding cause of death has been followed restrict process and the details can be found in the ONS mortality website(https://www.ons.gov.uk). We ascertained deaths by linking to mortality data and categorised causes of death using ICD-9 and ICD-10 codes to for the main underlying cause: CNS tumours, cardiovascular disease (CVD) and other causes. The primary outcome was death from CVD including diseases of heart, cerebrovascular disease, atherosclerosis, and aortic aneurysm or dissection(11).

##### SEER US

We used SEER Research Data, 18 Registries (excl AK) for SMRs for this analysis. Patients with malignant and non-malignant CNS tumours, diagnosed between 2005 and 2015, were abstracted from the National Cancer Institute's Surveillance, Epidemiology, and End Results (SEER) program, a network of population-based incident tumour registries from geographically distinct regions in the US, covering 28% of the US population, including incidence, survival, and treatment (12). The SEER registry does not code comorbidities, performance status, surgical pathology, margin status, doses, or agents. In SEER US, cause of death came from patient death certificates. The SEER database is representative of the population of the USA, and this has been validated by external studies(13-16).Mortality codes in SEER are assigned from death certificates, completed by the doctor caring for the patient at the time of demise. There could be some misclassification due to the coding of cause of death due to the index-cancer may not be perfect(17). Causes of deaths were coded as CNS tumours related death, cardiovascular disease, and other causes. SEER*Stat 8.3.8 was used for analysis(16). Patients diagnosed only through autopsy or death certificate were excluded.

#### Study population

We identified 8,743 patients diagnosed with CNS tumours between 2000 and 2015 from the WCISU data in SAIL (4,804 deaths; 54.9%) and 163,183 patients from SEER diagnosed between 2005 to 2015 (64,676 deaths; 39.6%). We sub-classified patients into those with malignant and with non-malignant primary CNS tumours using the following International Classification of Diseases 10$^{th}$ revision (ICD-10) codes: C70-72, C75.1-75.3, D18.0, D32-D33, D35.2-D35.4, D42-D43, D44.3-D44.5 (18)). We also used the International Classification of Diseases for Oncology third edition (ICD-O-3) codes to group tumours into malignancy type (malignant or non-malignant type) and three major histologic subgroups (meningiomas, gliomas, and all others), following the definitions from the Central Brain Tumour Registry of the United States (CBTRUS) (Online Appendix 1)(19). We restricted analysis to adults age 18 years or older based on the date of diagnosis.

#### Statistical analysis

Descriptive analyses of baseline characteristics by main causes of death in brain and CNS tumours were performed using the Pearson $\chi 2$ test for categorical variables (summarized as frequencies/percentages). Continuous variables were compared across subgroups using analysis of variance for normal distribution data presented as mean, 95% confidence interval (CI), or Kruskal–Wallis (summarized as medians and interquartile range) for non-normal distribution.

The first objective was to compare CVD mortality among patients with CNS tumours to the general population and expressed as standardised mortality ratios (SMRs). SMRs were calculated as the ratio of the observed to the expected number of death during study period, as measured in person-years, providing the relative risk of death for patients with CNS tumours as compared to the general population(20, 21).SMRs were calculated for the time period 2000-2015 for SAIL UK and 2005-2015 for SEER US. This is because non-malignant brain tumours were registered in SEER only from 2004 onwards(22). SMRs were adjusted by age, sex, calendar to the general population during the same time. The 95% confidence intervals (CIs) of the SMRs were calculated. Person-years were calculated from the date of study entry to the date of an event or end of study (31st December 2015). General population data stratified according to sex, age group and calendar period were used to calculate the expected number of events. Age was categorised and grouped 18-49 years due to low frequency of cases and 10-year-aged categories thereafter for SAIL and SEER. SMRs were estimated as a function of calendar year, age at cancer diagnosis, and follow-up time in those who survived more than 2 months after cancer diagnosis, respectively(11). We also performed SMR analysis for two main causes of death within CVD: heart disease and cerebrovascular disease.

Second object was to determine factors associated with elevated CVD mortality in CNS tumours patients. Survival analysis was performed and assessed by Kaplan-Meier methods and compared by the log-rank test to access all-cause and CVD-cause specific mortality. A competing risk analysis was applied by using multivariable cause-specific Cox proportional-hazard regression model to calculate the hazard ratios and 95% CI for CVD mortality as the primary outcome among different CNS tumours patient subgroups. For our study, deaths from other causes were censored at the time of death. The variables included in the Cox models were age at diagnosis, malignancy (malignant and non-malignant type), histological types (gliomas, meningioma, and other histological types), age at diagnosis, sex, race/ethnicity status (SEER), marital status (SEER) , area deprivation measured by Welsh Index of Multiple Deprivation (WIMD) (SAIL) and surgery status (having surgery vs No surgery/no information). Survival time was computed from the date of CNS cancer diagnosis and continued until date of death or last contact (December 31, 2015), and proportional hazards assumption was tested with the Schoenfeld residuals. We calculated cumulative incidence function curves to describe the incidence of deaths from CVD, and CNS tumours over time in all CNS tumour patients. Stratified analyses were also done by 1) malignant and non-malignant type; 2) major histological subtypes (glioma and meningioma, the most common malignant and non-malignant tumours, respectively, and by other histological types). We also performed analysis for all-cause mortality. To reduce the chance of reverse causality, all analyses were landmark analyses, with follow-up commencing 2 months after cancer diagnoses[16], thereby excluding patients with an event of death from cancer or CVD within 2 months (7). Additional analysis was performed for non-malignant CNS tumours 1 year after diagnosis. To test the robustness of our results and to address potential misclassification,  sensitivity analyses were performed in those who were diagnosed after 2005 when there has been stabilisation in incidence and diagnosis of CNS tumour patients in SAIL and histologically confirmed CNS tumours(18).

#### Part 2: Expanded materials

**Online Appendix 1.** International classification of diseases oncology version 3 (ICD-O-3) codes for tumour classification

**Online Appendix 2**. International classification of disease (ICD) code for cardiovascular mortality

**Online Appendix 3.** International Classification Codes for cause of CNS tumours

**Online Appendix 1. International classification of diseases oncology version 3 (ICD-O-3) codes for tumour classification(18)**

| Tumour categories | | ICD-O-3 codes* |
|---|---|---|
| **Meningioma** | | 9530, 9531, 9532, 9533, 9534, 9537, 9538, 9539, 8324, 8800, 8801, 8802, 8803, 8804, 8805, 8806, 8810, 8815, 8824, 8830, 8831, 8835, 8836, 8850, 8851, 8852, 8853, 8854, 8857, 8861, 8870, 8880, 8890, 8897, 8900, 8901, 8902, 8910, 8912, 8920, 8921, 8935, 8990, 9040, 9136, 9150, 9170, 9180, 9210, 9241, 9260, 9373, 8720, 8728, 8770, 8771, 9161, 9220, 9231, 9240, 9243, 9370, 9371, 9372, 9535 |
| **Glioma†** | **Grade I** | 93801, 9383, 9384, 9394, 9412, 9413, 9421, 9430, 9431 |
| | **Grade II** | 93931, 94000, 94001, 94003, 94103, 94110, 94113, 94203, 94243, 94303, 94441, 94500, 94501, 94503 |
| | **Grade III** | 93803, 93810, 93813, 93823, 93903, 9391, 9392, 94010, 94013, 94513, 94603, 9395 |
| | **Grade IV** | 94400, 94401, 94403, 94413, 94421, 94423 |
| **Others** | | 8680, 8681, 93611, 93900, 93901, 9492, 95051, 95050, 9509, 95053, 95060, 95061, 95063, 95223, 9540, 9560, 95710, 9590, 9591, 9596, 9650, 9651, 9652, 9653, 9654, 9655, 9659, 9661, 9662, 9663, 9664, 9665, 9667, 9670, 9671, 9673, 9675, 9680, 9684, 9687, 9690, 9691, 9695, 9698, 9699, 9701, 9702, 9705, 9714, 9719, 9728, 9729, 9727, 9731, 9733, 9734, 9740, 9741, 9750, 9751, 9752, 9753, 9754, 9755, 9756, 9757, 9758, 9760, 9766, 9823, 9826, 9827, 9832, 9837, 9860, 9861, 9866, 9930, 9970, 9360, 9361, 9362, 8963, 9364, 9470, 9471, 9472, 9473, 9474, 9480, 9490, 9500, 9501, 9502, 9508, 8020, 8440, 9060, 9061, 9064, 9065, 9070, 9071, 9072, 9080, 9081, 9082, 9083, 9084, 9085, 9100, 9101, 8040, 8140, 8146, 8246, 8260, 8270, 8271, 8272, 8280, 8281, 8290, 8300, 8310, 8323, 9492, 9582, 9120, 9121, 9122, 9123, 9125, 9130, 9131, 9133, 9140, 8000, 8001, 8002, 8003, 8004, 8005, 8010, 8021, 8320, 8452, 8710, 8711, 8713, 8811, 8840, 8896, 8980, 9173, 9503, 9580 |

*4-digit codes correspond to the histology codes; 5-digit codes correspond to the morphology codes, which is a combination of histology code and behaviour code.

†WHO grade according to Table 27 “WHO grading (malignancy scale) for central nervous system tumours” in the International Classification of Diseases Oncology, third edition, first revision (https://www.who.int/classifications/icd/adaptations/oncology/en/). When a code does not match with this reference table, we classify them based on literature search.

**Online Appendix 2. International Classification Codes for cardiovascular disease (11)**

| | ICD 10 | ICD 9 |
|---|---|---|
| **Cardiovascular disease** | | |
| **Disease of heart** | | |
| Acute rheumatic fever | I00-I02 | 390-392 |
| Chronic rheumatic heart disease | I05-I09 | 393-398 |
| Hypertensive disease | I10-I15 | 401-405 |
| Coronary artery disease (angina, myocardial infarction, revascularisation procedures) | I20-I 25 | 410-414 |
| Other forms of heart disease(sudden cardiac death, heart failure, pulmonary heart disease, | I30-I52 | 420—429 |
| **Cerebrovascular disease** | **I60-69** | **430-438** |
| Intracranial haemorrhagic | I60-I62 | 430, 431, 342 |
| Occlusive cerebrovascular accident | I63-68 | 433, 435, 436, 437 |
| Sequela of cerebrovascular disease | I69 | 438 |
| **Disease of arteries, arterioles and capillary**(including atherosclerosis, aortic aneurysm and dissection) | I70-I79 | 440-449 |

**Online Appendix 3. International Classification Codes for cancer cause of death**

| **Malignant brain and CNS tumours** | **ICD 10** | **ICD 9** |
|---|---|---|
| Malignant brain and CNS tumours | C70-72 | 191, 192, 194.3-194.4 |
| Malignant neoplasm of meninges | C70 | 192.1, 192.3 |
| Malignant neoplasm of brain | C71 | All 191 |
| Malignant neoplasm of spinal cord, cranial nerves and other parts of CNS | C72 | 192.0, 192.2, 192.8, 192.9 |
| Malignant of pituitary, pineal glands and craniopharyngeal duct | C75.1-C75.3 | 194.3, 1194.4 |
| **Non-malignant brain and CNS tumours** | **ICD 10** | **ICD 9** |
| Non-malignant brain and CNS tumours | D18.0, D32-D33, D35.2-D35.4, D42-D43, D44.3-D44.5 | 228, 225, 237 |
| Haemangioma | D180 | 228 |
| Benign neoplasm of meninges | D32 | 225.2, 225.4 |
| Benign neoplasm of brain and other part of CNS | D33 | 225.0, 225.1, 225.3, 225.8, 225.9 |
| Benign neoplasm t of pituitary, pineal glands and craniopharyngeal duct | D35.2-D35.4 | 227.3-227.4 |
| Neoplasm of uncertain or unknown behaviour of meninges | D42 | 237.6 |
| Neoplasm of uncertain or unknown behaviour of brain and CNS | D43 | 237.5, 237.6, 237.9 |
| Neoplasm of uncertain or unknown behaviour of endocrine glands | D44 | 237.0, 237.1 |

#### Part 3: Online Tables and Figures

**OnlineTable 1** Characteristics of data SAIL UK and SEER US

**OnlineTable 2** Cardiovascular mortality in CNS tumours by calendar year and stratified by subgroups (SAIL UK & SEER US)

**Online Table 3** Cardiovascular mortality in CNS tumours by age at diagnosis and stratified by subgroups (SAIL UK & SEER US)

**Online Table 4** SMRs for cardiovascular mortality by follow up time in CNS tumours and stratified by subgroups (SAIL UK & SEER US)

**Online Figure 1** Cumulative incidence function for cause-specific death among CNS tumours by main histological group (SAIL UK & SEER US)

**Online Table 1 Characteristics of SAIL UK and SEER US**

| Data | SAIL UK 2000-2015 | SEER US 2005-2015 |
|---|---|---|
| **Data sources** | Secure Anonymised Information Linkage (SAIL) Databank, including Welsh Cancer Registry, Mortality data, Welsh demographic data. Welsh Cancer registry has 100% representative of Welsh population | US National Cancer Institute's Surveillance, Epidemiology, and End Results (SEER) program, a network of cancer registries from geographic distinct region. SEER 18 Research data* is used, representing the largest geographic coverage available data 28% of the US population. |
| **Reference population** | Wales mortality data, UK | U.S. mortality data† |
| **Year** | Jan 2000 to Dec 2015 | Jan 2005 to Dec 2015 |
| **Selection of case** | CNS tumours | CNS tumours |
| **Primary selection** | | First primary in database |
| **Exclusion** | Age < 18 years | Age <18 years |
| **Variables for SMR‡ analysis** | | |
| **Patency exclusion period** | Survival months <=2 months | Survival months <=2 months |
| **Follow up years (period)** | <1 year, 1-4 year, 5-10 years, >=10 years | <1 year, 1-4 year, 5-10 years, >=10 years |
| **Age at diagnosis (years)** | 18-49, 50-59, 60-69, 70-79, 80+ years | 18-49 years, 50-59 years, 60-69 year, 70-79 years, 80+ years |
| **Attained calendar years** | 3-year interval | 2005, 2010, 2015 |
| **Malignancy** | Malignant and non-malignant types | Malignant and non-malignant types |
| **Histology main groups** | Glioma, meningioma and other histological types | Glioma, meningioma and other histological types |

**Footnotes: * Surveillance, Epidemiology, and End Results (SEER) Program (www.seer.cancer.gov) SEER*Stat Database: Incidence - SEER Research Data, 18 Registries (excl AK), Nov 2019 Sub (2000-2017) for SMRs - Linked To County Attributes - Time Dependent (1990-2017) Income/Rurality, 1969-2018 Counties, National Cancer Institute, DCCPS, Surveillance Research Program, released April 2020, based on the November 2019 submission. Largest geographic coverage available MP-SMRs- approximately 27.8% of the U.S. population (based on 2010 census); †Title: U.S. Mortality 1975-2017 (Nov 2019 sub), Race (WU/B/O), Event: COD rec (HIV grouped w/oth infectious); ‡SMR: Standardized mortality ratio**

**Table 2 Cardiovascular mortality in CNS tumours by calendar year and stratified by subgroups**

**SAIL UK 2000-2015**

| **Cardiovascular disease** | **Overall** | | | | | **Non-malignant type** | | | | | **Malignant type** | | | | |
|---|---|---|---|---|---|---|---|---|---|---|---|---|---|---|---|
| **Year** | **Person -years** | **SMR** | **Low** | **High** | **P value** | **Person-years** | **SMR** | **Low** | **High** | **P value** | **Person-years** | **SMR** | **Low** | **High** | **P value** |
| **2000-2003** | 1329 | 5.03 | 3.64 | 6.73 | 0.00 | 866 | 5.32 | 3.61 | 7.50 | 0.00 | 463 | 4.44 | 2.38 | 7.45 | 0.00 |
| **2004-2007** | 5315 | 2.56 | 2.03 | 3.19 | 0.00 | 4029 | 2.42 | 1.85 | 3.09 | 0.00 | 1286 | 3.20 | 1.94 | 4.91 | 0.00 |
| **2008-2011** | 9659 | 2.46 | 2.06 | 2.89 | 0.00 | 7760 | 2.57 | 2.14 | 3.06 | 0.00 | 1900 | 1.78 | 1.00 | 2.87 | 0.03 |
| **2012-2015** | 13830 | 2.53 | 2.18 | 2.93 | 0.00 | 11476 | 2.75 | 2.35 | 3.19 | 0.00 | 2355 | 1.16 | 0.60 | 1.99 | 0.62 |
| **Disease of heart** | **Overall** | | | | | **Non-malignant type** | | | | | **Malignant type** | | | | |
| **Year** | **Person -years** | **SMR** | **Low** | **High** | **P value** | **Person-years** | **SMR** | **Low** | **High** | **P value** | **Person-years** | **SMR** | **Low** | **High** | **P value** |
| **2000-2003** | 1329 | 3.80 | 2.50 | 5.49 | 0.00 | 866 | 3.63 | 2.13 | 5.70 | 0.00 | 463 | 4.15 | 1.99 | 7.48 | 0.00 |
| **2004-2007** | 5315 | 2.04 | 1.48 | 2.73 | 0.00 | 4029 | 2.07 | 1.45 | 2.85 | 0.00 | 1286 | 1.90 | 0.82 | 3.67 | 0.09 |
| **2008-2011** | 9659 | 2.17 | 1.71 | 2.69 | 0.00 | 7760 | 2.19 | 1.70 | 2.76 | 0.00 | 1900 | 2.01 | 1.01 | 3.52 | 0.03 |
| **2012-2015** | 13830 | 2.10 | 1.67 | 2.60 | 0.00 | 11476 | 2.31 | 1.83 | 2.86 | 0.00 | 2355 | 0.78 | 0.24 | 1.81 | 0.62 |
| **Cerebrovascular disease** | **Overall** | | | | | **Non-malignant type** | | | | | **Malignant type** | | | | |
| **Year** | **Person -years** | **SMR** | **Low** | **High** | **P value** | **Person-years** | **SMR** | **Low** | **High** | **P value** | **Person-years** | **SMR** | **Low** | **High** | **P value** |
| **2000-2003** | 1329 | 10.76 | 6.06 | 17.42 | 0.00 | 866 | 12.48 | 6.48 | 21.37 | 0.00 | 463 | 7.16 | 1.78 | 18.55 | 0.00 |
| **2004-2007** | 5315 | 6.35 | 4.37 | 8.86 | 0.00 | 4029 | 5.44 | 3.47 | 8.05 | 0.00 | 1286 | 10.71 | 5.15 | 19.30 | 0.00 |
| **2008-2011** | 9659 | 6.06 | 4.57 | 7.84 | 0.00 | 7760 | 6.44 | 4.80 | 8.42 | 0.00 | 1900 | 3.49 | 1.08 | 8.11 | 0.01 |

| **2012-2015** | 13830 | 9.73 | 7.79 | 11.96 | 0.00 | 11476 | 10.22 | 8.11 | 12.68 | 0.00 | 2355 | 6.34 | 2.72 | 12.26 | 0.00 |
|---|---|---|---|---|---|---|---|---|---|---|---|---|---|---|---|

**Standardized mortality ratios (SMR) for cardiovascular disease (CVD) adjusted by age, sex in CNS tumours, and by malignancy subtypes. Risk for subtypes within cause of CVD (disease of heart and cerebrovascular disease) were presented. A SMR above 1 represents a higher relative risk of death for a type of cardiovascular cause compared with the general population.**

**SEER US 2005-2015**

| **Cardiovascular disease** | | **Overall** | | | | **Non-malignant type** | | | | **Malignant type** | | |
|---|---|---|---|---|---|---|---|---|---|---|---|---|
| **Year** | **Person-years** | **SMR** | **Low** | **High** | **Person-years** | **SMR** | **Low** | **High** | **Person-years** | **SMR** | **Low** | **High** |
| **2005-2009** | 183994 | 1.53* | 1.46 | 1.61 | 115546 | 1.38* | 1.31 | 1.46 | 68448 | 2.49* | 2.24 | 2.77 |
| **2010-2014** | 397311 | 1.33* | 1.29 | 1.38 | 295308 | 1.27* | 1.23 | 1.32 | 102002 | 2.03* | 1.84 | 2.24 |
| **2015+** | 103133 | 1.32* | 1.23 | 1.41 | 79599 | 1.26* | 1.18 | 1.35 | 23534 | 2.08* | 1.69 | 2.55 |
| **Diseases of Heart** | | **Overall** | | | | **Non-malignant type** | | | | **Malignant type** | | |
| **Year** | **Person-years** | **SMR** | **Low** | **High** | **Person-years** | **SMR** | **Low** | **High** | **Person-years** | **SMR** | **Low** | **High** |
| **2005-2009** | 183994 | 1.40* | 1.32 | 1.48 | 115546 | 1.25* | 1.17 | 1.34 | 68448 | 2.32* | 2.04 | 2.62 |
| **2010-2014** | 397311 | 1.23* | 1.18 | 1.28 | 295308 | 1.19* | 1.13 | 1.24 | 102002 | 1.73* | 1.53 | 1.95 |
| **2015+** | 103133 | 1.18* | 1.09 | 1.28 | 79599 | 1.15* | 1.05 | 1.25 | 23534 | 1.61* | 1.22 | 2.09 |
| **Cerebrovascular Diseases** | | **Overall** | | | | **Non-malignant type** | | | | **Malignant type** | | |
| **Year** | **Person-years** | **SMR** | **Low** | **High** | **Person-years** | **SMR** | **Low** | **High** | **Person-years** | **SMR** | **Low** | **High** |
| **2005-2009** | 183994 | 2.16* | 1.96 | 2.38 | 115546 | 1.99* | 1.78 | 2.21 | 68448 | 3.46* | 2.74 | 4.32 |
| **2010-2014** | 397311 | 1.74* | 1.62 | 1.87 | 295308 | 1.61* | 1.48 | 1.73 | 102002 | 3.53* | 2.9 | 4.27 |
| **2015+** | 103133 | 1.81* | 1.58 | 2.06 | 79599 | 1.65* | 1.43 | 1.91 | 23534 | 4.32* | 2.94 | 6.13 |

**Standardized mortality ratios (SMR) for cardiovascular disease (CVD) adjusted by age, sex in total CNS tumours, and by malignancy subtypes. Risk for subtypes within cause of CVD (disease of heart and cerebrovascular disease) were presented. A SMR above 1 represents a higher relative risk of death for a type of cardiovascular cause compared with the general population. * p<0.05**

**Online Table 3 Cardiovascular mortality in CNS tumours by age at diagnosis and stratified by subgroups SAIL UK 2000-2015**

| **Cardiovascular disease** | **Overall** | | | | | **Non-malignant type** | | | | | **Malignant type** | | | | |
|---|---|---|---|---|---|---|---|---|---|---|---|---|---|---|---|
| **Age group** | **Person-years** | **SMR** | **Low** | **High** | **P value** | **Person-years** | **SMR** | **Low** | **High** | **P value** | **Person-years** | **SMR** | **Low** | **High** | **P value** |
| **18-49 years** | 10146 | 96.09 | 48.89 | 164.55 | 0.000 | 7600 | 115.4 | 55.45 | 208 | 0.000 | 2546 | 54.82 | 9.11 | 169.16 | 0.000 |
| **50-59 years** | 5790 | 7.32 | 4.94 | 10.38 | 0.000 | 4642 | 6.7 | 4.17 | 10.08 | 0.000 | 1148 | 9.56 | 4.37 | 17.79 | 0.000 |
| **60-69 years** | 6471 | 3.7 | 2.68 | 4.96 | 0.000 | 5316 | 3.93 | 2.77 | 5.38 | 0.000 | 1155 | 2.78 | 1.1 | 5.63 | 0.012 |
| **70-79 years** | 4651 | 4.33 | 3.52 | 5.25 | 0.000 | 3893 | 4.36 | 3.47 | 5.4 | 0.000 | 758 | 4.16 | 2.48 | 6.47 | 0.000 |
| **80+ years** | 3076 | 5.1 | 4.5 | 5.76 | 0.000 | 2680 | 5.41 | 4.74 | 6.14 | 0.000 | 396 | 3.21 | 2.05 | 4.75 | 0.000 |
| **Disease of heart** | **Overall** | | | | | **Non-malignant type** | | | | | **Malignant type** | | | | |
| **Age group** | **Person-years** | **SMR** | **Low** | **High** | **P value** | **Person-years** | **SMR** | **Low** | **High** | **P value** | **Person-years** | **SMR** | **Low** | **High** | **P value** |
| **18-49 years** | 10146 | 65.17 | 20.23 | 151.36 | 0.000 | 7600 | 71.47 | 17.77 | 185.25 | 0.000 | 2546 | 51.54 | 2.94 | 226.69 | 0.001 |
| **50-59 years** | 5790 | 6.04 | 3.6 | 9.38 | 0.000 | 4642 | 5.04 | 2.62 | 8.63 | 0.000 | 1148 | 9.49 | 3.77 | 19.22 | 0.000 |
| **60-69 years** | 6471 | 2.81 | 1.83 | 4.09 | 0.001 | 5316 | 2.91 | 1.82 | 4.38 | 0.000 | 1155 | 2.39 | 0.74 | 5.56 | 0.081 |
| **70-79 years** | 4651 | 3.96 | 3.06 | 5.02 | 0.000 | 3893 | 4.01 | 3.02 | 5.21 | 0.000 | 758 | 3.72 | 1.93 | 6.37 | 0.000 |
| **80+ years** | 3076 | 3.6 | 2.98 | 4.3 | 0.000 | 2680 | 3.89 | 3.2 | 4.68 | 0.000 | 396 | 1.8 | 0.82 | 3.36 | 0.096 |
| **Cerebrovascular disease** | **Overall** | | | | | **Non-malignant type** | | | | | **Malignant type** | | | | |
| **Age group** | **Person-years** | **SMR** | **Low** | **High** | **p value** | **Person-years** | **SMR** | **Low** | **High** | **p value** | **Person-years** | **SMR** | **Low** | **High** | **p value** |

| | | | | | | | | | | | | | | | |
|---|---|---|---|---|---|---|---|---|---|---|---|---|---|---|---|
| **18-49 years** | 10146 | 321.97 | 138.34 | 622.62 | 0.000 | 7600 | 456.73 | 181.52 | 925.42 | 0.000 | 2546 | 116.22 | 6.33 | 511.23 | 0.000 |
| **50-59 years** | 5790 | 10.68 | 5.35 | 18.73 | 0.000 | 4642 | 10.68 | 4.88 | 19.78 | 0.000 | 1148 | 10.7 | 1.78 | 33 | 0.001 |
| **60-69 years** | 6471 | 5.6 | 3 | 9.39 | 0.000 | 5316 | 5.76 | 2.89 | 10.11 | 0.000 | 1155 | 4.91 | 0.82 | 15.14 | 0.025 |
| **70-79 years** | 4651 | 6.11 | 4.11 | 8.66 | 0.000 | 3893 | 5.84 | 3.73 | 8.64 | 0.000 | 758 | 7.3 | 2.9 | 14.8 | 0.000 |
| **80+ years** | 3076 | 8.56 | 7.14 | 10.15 | 0.000 | 2680 | 8.95 | 7.4 | 10.7 | 0.000 | 396 | 6.07 | 3.25 | 10.18 | 0.000 |

**Standardized mortality ratios (SMR) for cardiovascular disease (CVD) adjusted by age, sex in total CNS tumours, and by malignancy subtypes. Risk for subtypes within cause of CVD (disease of heart and cerebrovascular disease) were presented. A SMR above 1 represents a higher relative risk of death for a type of cardiovascular cause compared with the general population.**

**SEER US 2005-2015**

| **Cardiovascular disease** | **Overall** | | | | **Non-malignant type** | | | | **Malignant type** | | | |
|---|---|---|---|---|---|---|---|---|---|---|---|---|
| **Age group** | **Person-years** | **SMR** | **Low** | **High** | **Person-years** | **SMR** | **Low** | **High** | **Person-years** | **SMR** | **Low** | **High** |
| **18-49 years** | 225871 | 2.03* | 1.79 | 2.30 | 140102 | 1.82* | 1.53 | 2.14 | 857670 | 2.41* | 1.98 | 2.91 |
| **50-59 years** | 138817 | 1.64* | 1.5 | 1.80 | 111722 | 1.39* | 1.24 | 1.56 | 27095 | 2.56* | 2.17 | 2.99 |
| **60-69 years** | 119273 | 1.45* | 1.35 | 1.56 | 102138 | 1.28* | 1.18 | 1.39 | 17135 | 2.43* | 2.1 | 2.79 |
| **70-79 years** | 827780 | 1.30* | 1.23 | 1.36 | 75173 | 1.21* | 1.14 | 1.27 | 7607 | 2.22* | 1.94 | 2.54 |
| **80+ years** | 46770 | 1.33* | 1.28 | 1.38 | 44460 | 1.32* | 1.27 | 1.37 | 2310 | 1.59* | 1.35 | 1.87 |
| **Disease of heart** | **Overall** | | | | **Non-malignant type** | | | | **Malignant type** | | | |
| **Age group** | **Person-years** | **SMR** | **Low** | **High** | **Person-years** | **SMR** | **Low** | **High** | **Person-years** | **SMR** | **Low** | **High** |
| **18-49 years** | 225871 | 1.55* | 1.31 | 1.81 | 140102 | 1.49* | 1.21 | 1.82 | 857670 | 1.64* | 1.25 | 2.11 |

| | | | | | | | | | | | |
|---|---|---|---|---|---|---|---|---|---|---|---|
| **50-59 years** | 138817 | 1.45* | 1.29 | 1.62 | 111722 | 1.24* | 1.08 | 1.41 | 27095 | 2.20* | 1.8 | 2.65 |
| **60-69 years** | 119273 | 1.31* | 1.2 | 1.43 | 102138 | 1.16* | 1.05 | 1.28 | 17135 | 2.16* | 1.81 | 2.55 |
| **70-79 years** | 827780 | 1.20* | 1.13 | 1.27 | 75173 | 1.11* | 1.03 | 1.18 | 7607 | 2.12* | 1.81 | 2.47 |
| **80+ years** | 46770 | 1.24* | 1.19 | 1.3 | 44460 | 1.23* | 1.18 | 1.29 | 2310 | 1.47* | 1.2 | 1.78 |
| **Cerebrovascular disease** | **Overall** | | | | **Non-malignant type** | | | | **Malignant type** | | | |
| **Age group** | **Person-years** | **SMR** | **Low** | **High** | **Person-years** | **SMR** | **Low** | **High** | **Person-years** | **SMR** | **Low** | **High** |
| **18-49 years** | 225871 | 4.82* | 3.84 | 5.99 | 140102 | 3.42* | 2.44 | 4.68 | 85770 | 7.66* | 5.54 | 10.31 |
| **50-59 years** | 138817 | 2.86* | 2.34 | 3.45 | 111722 | 2.28* | 1.77 | 2.88 | 27095 | 5.19* | 3.69 | 7.09 |
| **60-69 years** | 119273 | 2.27* | 1.96 | 2.61 | 102138 | 1.98* | 1.67 | 2.33 | 17135 | 4.05* | 2.99 | 5.37 |
| **70-79 years** | 82780 | 1.65* | 1.48 | 1.83 | 75173 | 1.55* | 1.38 | 1.73 | 7607 | 2.76* | 2.05 | 3.66 |
| **80+ years** | 46770 | 1.64* | 1.51 | 1.77 | 44460 | 1.63* | 1.5 | 1.76 | 2310 | 1.76* | 1.18 | 2.51 |

**Standardized mortality ratios (SMR) for cardiovascular disease (CVD) adjusted by age, sex in total CNS tumours, and by malignancy subtypes. Risk for subtypes within cause of CVD (disease of heart and cerebrovascular disease) were presented. A SMR above 1 represents a higher relative risk of death for a type of cardiovascular cause compared with the general population. * <0.05**

**Online Table 4 Cardiovascular mortality by follow up time in CNS tumours and stratified by subgroups SAIL UK 2000-2015**

| **Cardiovascular death** | **Overall** | | | | | **Non-malignant type** | | | | | **Malignant type** | | | | |
|---|---|---|---|---|---|---|---|---|---|---|---|---|---|---|---|
| **Follow up time** | **Person-years** | **SMR** | **Low** | **High** | **p value** | **Person-years** | **SMR** | **Low** | **High** | **p value** | **Person-years** | **SMR** | **Low** | **High** | **P value** |
| **2-11 months** | 4861 | 2.98 | 2.39 | 3.66 | 0.00 | 3404 | 3.38 | 2.67 | 4.22 | 0.00 | 1456 | 1.72 | 0.92 | 2.88 | 0.06 |

| | | | | | | | | | | | | | | | |
|---|---|---|---|---|---|---|---|---|---|---|---|---|---|---|---|
| **1-4 years** | 13816 | 2.18 | 1.86 | 2.54 | 0.00 | 11310 | 2.31 | 1.96 | 2.70 | 0.00 | 2507 | 1.23 | 0.64 | 2.11 | 0.49 |
| **5-9 years** | 8056 | 1.68 | 1.31 | 2.11 | 0.00 | 6867 | 1.74 | 1.34 | 2.21 | 0.00 | 1190 | 1.18 | 0.42 | 2.54 | 0.71 |
| **10+ years** | 2062 | 1.36 | 0.78 | 2.17 | 0.23 | 1749 | 1.26 | 0.67 | 2.11 | 0.43 | 313 | 2.03 | 0.50 | 5.26 | 0.22 |
| **Heart disease** | **Overall** | | | | | **Non-malignant type** | | | | | **Malignant type** | | | | |
| **Follow up time** | **Person-years** | **SMR** | **Low** | **High** | **p value** | **Person-years** | **SMR** | **Low** | **High** | **p value** | **Person-years** | **SMR** | **Low** | **High** | **p value** |
| **2-11 months** | 4861 | 2.68 | 1.98 | 3.53 | 0.00 | 3404 | 2.85 | 2.03 | 3.87 | 0.00 | 1456 | 2.14 | 1.11 | 4.11 | 0.02 |
| **1-4 years** | 13816 | 2.08 | 1.68 | 2.55 | 0.00 | 11310 | 2.24 | 1.79 | 2.76 | 0.00 | 2507 | 0.98 | 0.41 | 2.35 | 0.96 |
| **5-9 years** | 8056 | 1.69 | 1.19 | 2.30 | 0.00 | 6867 | 1.83 | 1.29 | 2.51 | 0.00 | 1190 | 0.44 | 0.06 | 3.16 | 0.42 |
| **10+ years** | 2062 | 0.87 | 0.31 | 1.86 | 0.75 | 1749 | 1.00 | 0.36 | 2.15 | 1.00 | 313 | 0.00 | 0.00 | Inf | 1.00 |
| **Cerebrovascular disease** | **Overall** | | | | | **Non-malignant type** | | | | | **Malignant type** | | | | |
| **Follow up time** | **Person-years** | **SMR** | **Low** | **High** | **p value** | **Person-years** | **SMR** | **Low** | **High** | **p value** | **Person-years** | **SMR** | **Low** | **High** | **p value** |
| **2-11 months** | 4861 | 7.85 | 5.46 | 10.84 | 0.00 | 3404 | 9.13 | 6.24 | 12.79 | 0.00 | 1456 | 3.27 | 0.81 | 8.47 | 0.04 |
| **1-4 years** | 13816 | 6.21 | 4.81 | 7.86 | 0.00 | 11310 | 6.59 | 5.06 | 8.41 | 0.00 | 2507 | 3.32 | 1.03 | 7.72 | 0.02 |

| | | | | | | | | | | | | | | |
|---|---|---|---|---|---|---|---|---|---|---|---|---|---|---|
| **5-9 years** | 8056 | 5.19 | 3.47 | 7.40 | 0.00 | 6867 | 4.95 | 3.19 | 7.26 | 0.00 | 1190 | 7.15 | 2.22 | 16.60 | 0.00 |
| **10+ years** | 2062 | 6.48 | 3.12 | 11.69 | 0.00 | 1749 | 5.03 | 2.00 | 10.18 | 0.00 | 313 | 15.45 | 3.84 | 40.04 | 0.00 |

**Standardized mortality ratios (SMR) for cardiovascular disease (CVD) adjusted by age, sex in total CNS tumours, and by malignancy subtypes. Risk for subtypes within cause of CVD (disease of heart and cerebrovascular disease) were presented. A SMR above 1 represents a higher relative risk of death for a type of cardiovascular cause compared with the general population.**

**SEER US 2005-2015**

| **Cardiovascular disease** | **Overall** | | | | **Non-malignant type** | | | | **Malignant type** | | | |
|---|---|---|---|---|---|---|---|---|---|---|---|---|
| **Follow up time** | **Person-years** | **SMR** | **Low** | **High** | **Person-years** | **SMR** | **Low** | **High** | **Person-years** | **SMR** | **Low** | **High** |
| **2-11 months** | 109901 | 2.14* | 2.03 | 2.25 | 77308 | 1.92* | 1.81 | 2.03 | 32593 | 3.41* | 3.06 | 3.78 |
| **1-4 years** | 352800 | 1.25* | 1.21 | 1.3 | 268225 | 1.19* | 1.15 | 1.24 | 84575 | 2.04* | 1.82 | 2.28 |
| **5-9 years** | 192163 | 1.20* | 1.13 | 1.26 | 135087 | 1.18* | 1.12 | 1.25 | 57077 | 1.39* | 1.16 | 1.66 |
| **10+ years** | 29573 | 1.17 | 0.98 | 1.39 | 9834 | 1.03 | 0.82 | 1.28 | 19740 | 1.50* | 1.12 | 1.97 |
| **Disease of heart** | **Overall** | | | | **Non-malignant type** | | | | **Malignant type** | | | |
| **Follow up time** | **Person-years** | **SMR** | **Low** | **High** | **Person-years** | **SMR** | **Low** | **High** | **Person-years** | **SMR** | **Low** | **High** |
| **2-11 months** | 109901 | 1.94* | 1.82 | 2.06 | 77308 | 1.70* | 1.58 | 1.82 | 32593 | 3.24* | 2.86 | 3.65 |
| **1-4 years** | 352799 | 1.15* | 1.10 | 1.20 | 268225 | 1.10* | 1.05 | 1.16 | 84575 | 1.72* | 1.50 | 1.97 |

| | | | | | | | | | | | | |
|---|---|---|---|---|---|---|---|---|---|---|---|---|
| **5-9 years** | 192163 | 1.11* | 1.04 | 1.19 | 135087 | 1.11* | 1.04 | 1.19 | 57077 | 1.09 | 0.86 | 1.36 |
| **10+ years** | 29573 | 1.05 | 0.84 | 1.29 | 9834 | 0.98 | 0.75 | 1.27 | 19740 | 1.20 | 0.82 | 1.69 |
| **Cerebrovascular disease** | **Overall** | | | | **Non-malignant type** | | | | **Malignant type** | | | |
| **Follow up time** | **Person-years** | **SMR** | **Low** | **High** | **Person-years** | **SMR** | **Low** | **High** | **Person-years** | **SMR** | **Low** | **High** |
| **2-11 months** | 109901 | 3.05* | 2.75 | 3.38 | 77308 | 2.92* | 2.6 | 3.26 | 32593 | 3.92* | 3.02 | 5.00 |
| **1-4 years** | 352799 | 1.67* | 1.55 | 1.80 | 268225 | 1.53* | 1.41 | 1.66 | 84575 | 3.90* | 3.15 | 4.77 |
| **5-9 years** | 192163 | 1.54* | 1.37 | 1.72 | 135087 | 1.43* | 1.26 | 1.61 | 57077 | 3.00* | 2.17 | 4.05 |
| **10+ years** | 29573 | 1.59* | 1.09 | 2.24 | 9834 | 1.09 | 0.62 | 1.77 | 19740 | 2.92* | 1.67 | 4.75 |

**Standardized mortality ratios (SMR) for cardiovascular disease (CVD) adjusted by age, sex in total CNS tumours, and by malignancy subtypes. Risk for subtypes within cause of CVD (disease of heart and cerebrovascular disease) were presented. A SMR above 1 represents a higher relative risk of death for a type of cardiovascular cause compared with the general population. * p<0.05**

**Online Table 5 Cox proportional hazards regression multivariable HRs and 95% CIs for associations between patient characteristics and all-cause and CVD-mortality**

**SAIL UK 2000-2015**

| | **All-cause mortality** | **Cardiovascular disease mortality deaths** | | |
|---|---|---|---|---|
| | **All-cause mortality** | **Overall** | **Malignancy** | |
| | | | **Non-malignant type** | **Malignant type** |
| | | **CVD mortality** | **CVD mortality** | **CVD mortality** |

| Characteristic | HR(95% CI) | HR(95% CI) | HR(95% CI) | HR(95% CI) |
|---|---|---|---|---|
| **Malignancy** | | | | |
| **Malignant type** | 1 | 1 | | |
| **Non-malignant type** | 0.32(0.29-0.35)* | 1.82(1.16-2.84) † | | |
| **Histology type** | | | | |
| **Gliomas** | 1 | 1 | 1 | 1 |
| **Meningioma** | 0.48(0.42-0.55)* | 1.29(0.74-2.23) | 1.24(0.31-5.04) | 0.72(0.17-3.05) |
| **Other** | 0.49(0.44-0.54)* | 1.11(0.66-1.88) | 1.08(0.27-4.39) | 1.32(0.69-2.52) |
| **Gender** | | | | |
| **Female** | 1 | 1 | 1 | 1 |
| **Male** | 1.13(1.06-1.20)* | 1.23(1.01-1.50)* | 1.17(0.95-1.45) | 1.73(0.99-3.02) |
| **Age** | 1.06(1.05-1.06)* | 1.10(1.09-1.11)* | 1.10(1.09-1.11)* | 1.08(1.05-1.10)* |
| **Area deprivation** | | | | |
| **WIMD 5(Least deprived)** | 1 | 1 | 1 | 1 |
| **4** | 1.02(0.94-1.11) | 0.95(0.69-1.30) | 1.00(0.71-1.40) | 0.62(0.26-1.47) |
| **3** | 1.04(0.95-1.14) | 1.32(0.98-1.77) | 1.34(0.97-1.85) | 1.05(0.49-2.25) |
| **2** | 1.14(1.04-1.34)** | 1.26(0.93-1.71) | 1.40(1.01-1.94) † | 0.50(0.2-1.29) |
| **1 Most deprived** | 1.25(1.14-1.37)* | 1.75(1.30-2.35)* | 1.85(1.35-2.55)* | 1.03(0.45-2.37) |
| **Calendar year** | 1.03(0.95-1.12) | 0.98(0.96-1.01) | 0.99(0.96-1.11) | 0.91(0.84-0.99) |
| **Treatment** | | | | |
| **Treatment** | 1 | 1 | 1 | 1 |

| No information on treatment | 1.35(1.27-1.45)* | 1.43(1.15-1.79)* | 1.43(1.15-1.79)* | 1.85(0.96-3.57) |
|---|---|---|---|---|

**HR: Hazard ratio; CI: Confidence interval; CVD: Cardiovascular disease, WIMD: Welsh Index Multiple Deprivation; *<0.001, ** <0.01; *<0.05**

**SEER US 2005-2015**

| | **All-cause mortality** | **Cardiovascular disease mortality** | | |
|---|---|---|---|---|
| | **All-cause mortality** | **Overall** | **Malignancy** | |
| | | | **Non-malignant type** | **Malignant type** |
| | | **CVD mortality** | **CVD mortality** | **CVD mortality** |
| **Characteristics** | **HR(95% CI)** | **HR(95% CI)** | **HR(95% CI)** | **HR(95% CI)** |
| **Malignancy** | | | | |
| **Malignant type** | 1.00 | 1.00 | | |
| **Non-malignant type** | 0.15(0.14-0.15)* | 0.56(0.49-0.64)* | | |
| **Histology type** | | | | |
| **Gliomas** | 1.00 | 1.00 | 1.00 | 1.00 |
| **Meningioma** | 0.59(0.56-0.61)* | 1.43(1.23-1.67)* | 1.52(1.00-2.32) ‡ | 1.17(0.90-1.52) |
| **Other** | 0.56(0.54-0.58)* | 1.03(0.88-1.19) | 1.09(0.71-1.67) | 1.36(1.13-1.65) † |
| **Gender** | | | | |
| **Female** | 1.00 | 1.00 | 1.00 | 1.00 |
| **Male** | 1.29(1.27-1.31)* | 1.61(1.53-1.69)* | 1.64(1.55-1.74)* | 1.38(1.22-1.57)* |
| **Age** | 1.06(1.06-1.06)* | 1.09(1.09-1.10)* | 1.10(1.10-1.10)* | 1.07(1.07-1.08)* |
| **Ethnicity** | | | | |
| **Non-Hispanic White** | 1.00 | | 1.00 | 1.00 |
| **Hispanic (All races)** | 1.00(0.97-1.03) | 0.98(0.90-1.07) | 0.95(0.87-1.05) | 1.13(0.92-1.39) |
| **Non-Hispanic Black** | 1.15(1.12-1.19)* | 1.55(1.44-1.66)* | 1.53(1.42-1.65)* | 1.78(1.45-2.18)* |
| **Non-Hispanic Asian or Pacific Islander** | 0.86(0.83-0.89)* | 0.84(0.76-0.93) | 0.83(0.75-0.92)* | 0.95(0.71-1.25) |
| **Non-Hispanic American Indian/Alaska Native** | 0.95(0.84-1.07) | 0.94(0.66-1.35) | 0.84(0.56-1.27) | 1.63(0.77-3.44) |
| **Non-Hispanic Unknow race** | 0.24(0.19-0.29)* | 0.18(0.09-0.38) | 0.17(0.08-0.38)* | 0.34(0.05-2.40) |

| | | | | |
|---|---|---|---|---|
| **Marital status** | | | | |
| **Married/Having Partner** | 1.00 | 1.00 | 1.00 | 1.00 |
| **Single/Separate/Divorced** | 1.43(1.40-1.46)* | 1.45(1.38-1.53)* | 1.42(1.34-1.50)* | 1.47(1.29-1.67)* |
| **Unknown** | 0.99(0.95-1.00) | 1.25(1.14-1.37)* | 1.25(1.13-1.38)* | 1.13(0.84-1.54) |
| **Year of diagnosis** | 0.99(0.99-1.00)* | 0.98(0.98-0.99)* | 0.99(0.98-1.00)* | 0.98(0.94-0.98)* |
| **Treatment** | | | | |
| **Surgery** | 1.00 | 1.00 | 1.00 | 1.00 |
| **No surgery or no information on surgery** | 1.47(1.45-1.49)* | 1.34(1.27-1.43)* | 1.30(1.22-1.37)* | 1.56(1.37-1.79)* |

HR: Hazard ratio; CI: Confidence interval; CVD: Cardiovascular disease. ***<0.001, †<0.01; ‡<0.05

**Online Figure 1 Cumulative incidence function for cause-specific death among CNS tumours by main histological groups**

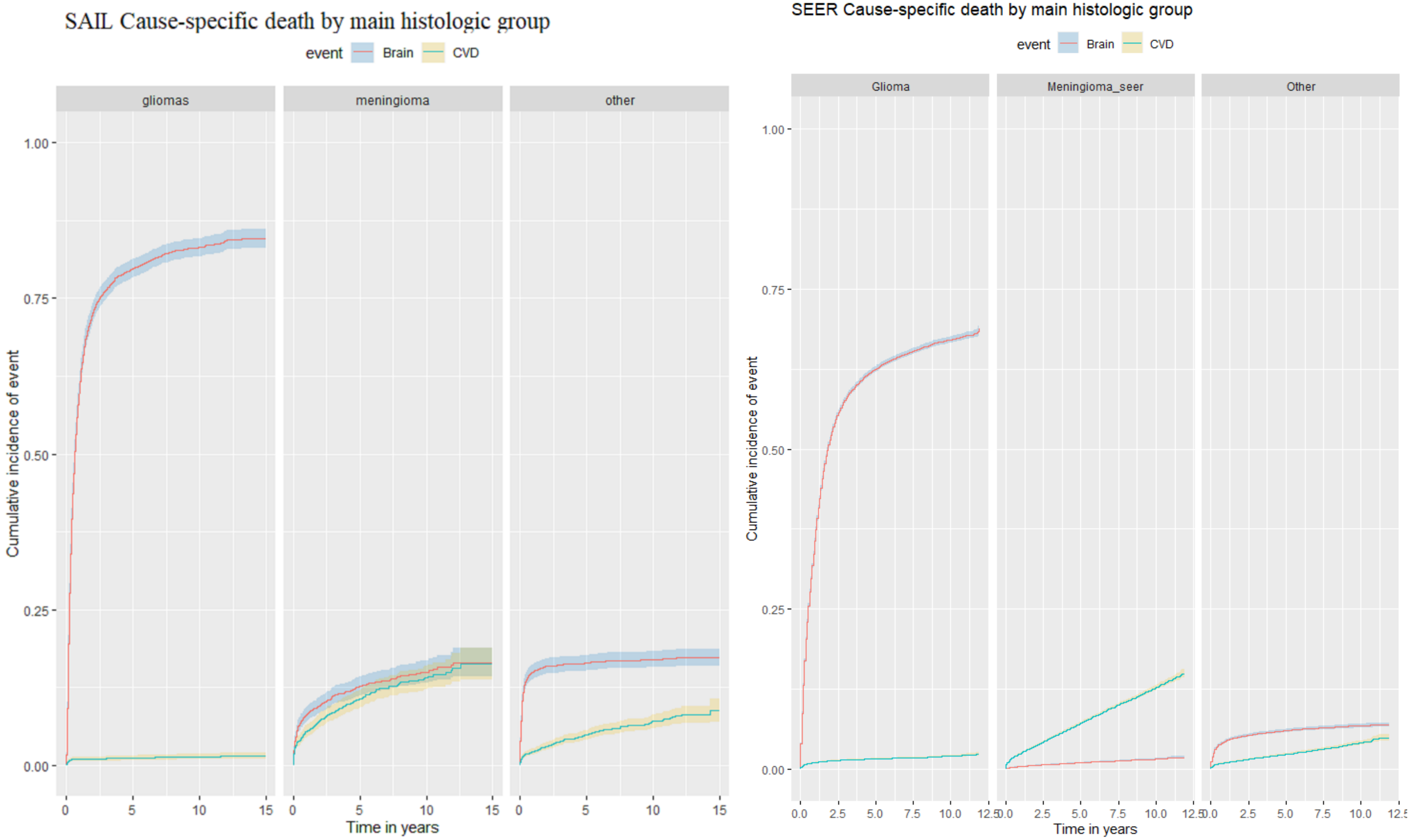

### Part 4: Systematic review

#### Systematic review of previous studies comparing cardiovascular mortality in central nervous system tumours patients with the general population

##### Methods

**Databases searched:**
OVID Medline & EMBASE

**Search strategies**
A full title search (search 1) was combined with a broader title/abstract search of key journals (search 2) as below

**Key words**
**Brain and central nervous system (CNS) tumours related terms:**

1. Brain neoplasm
2. Neuro-oncology
3. Gliomas/glioblastoma/astrocytoma/meningioma/schwannoma/oliodendroglioma/medulloblastoma/ependymoma
4. Intracranial tumour/cancers
5. Brain tumour or brain cancer or central nervous system tumour(s)

1 OR 2 OR 3 OR 4 OR 5

**Cardiovascular outcomes:**

Cardiovascular disease

Heart disease:

Acute Myocardial Infarction (STEMI-elevation MI, non-STEMI elevation MI) – includes or excludes post-MI revascularization; medically managed Coronary Artery Disease (CAD) – (e.g. stable angina); Revascularization procedures including percutaneous coronary interventions and/or coronary artery bypass graft surgery,

Heart failure

AF, ventricular arrhythmias

Stroke, cerebrovascular,

Venous thrombosis

Peripheral vascular disease

**Study types**
Epidemiologist studies, cohort, population based, case-control including clinical trials, systematic reviews, cohort studies, cross-sectional studies, observational studies, interventional studies, case series, and pilot studies.

**Period**
English language Jan 1998-2021 Jan

Additionally, the search included reference list, manual search from google and grey literature, review paper.

**Inclusion criteria**
Articles were included in the full text screen if they provided relative estimates comparing risk of cardiovascular disease mortality in adult CNS tumours survivors with cancer-free controls.

**Exclusion criteria**
Articles were excluded if they did not report estimates for CNS cancers and specific cardiovascular diseases included in our study.

**Extraction of study characteristics**
The following were extracted from each included study where information was available: data source /setting, countries, brain tumour site(s), compared group, age profile, specific cardiovascular outcomes investigated, adjustment for covariates, average follow-up.

**Extraction of study results**
Standardised mortality ratios or hazard ratios were extracted directly.

**Processing of study results**
Descriptive characteristics of the included studies are presented in table form. Results of all studies are summarised graphically.

**Flowchart**
Review of previous studies: flow chart of the systematic review search process

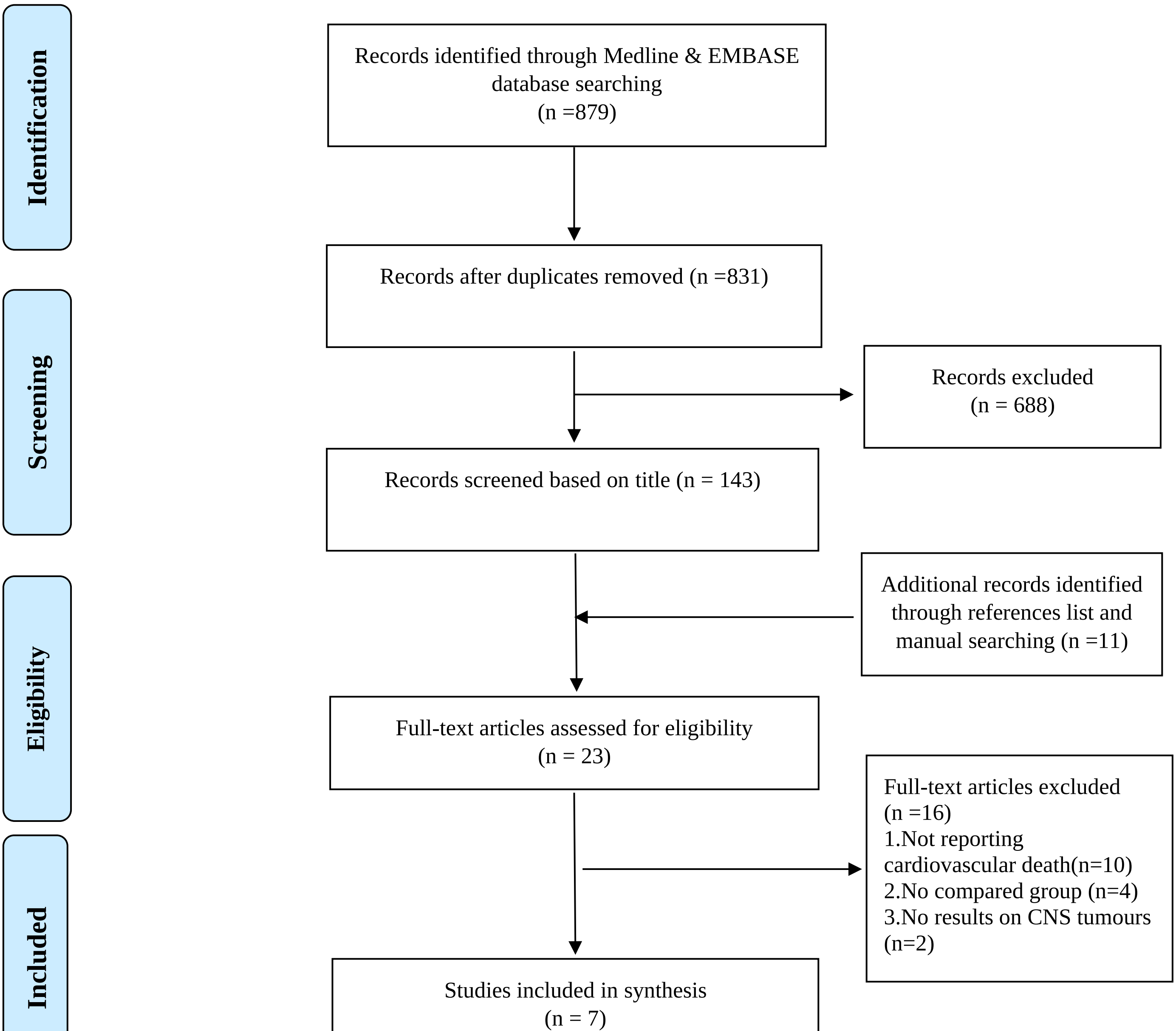

**Table 1 Summary of the results**

| Authors/ year | Country | Data source/Setting | Cancer sites | Period | Follow up | Study size | Age profile | Compared group | Outcome Specific CVD mortality | Adjustment for covariates |
|---|---|---|---|---|---|---|---|---|---|---|
| **Brada et al 2002(1)** | UK | Following surgery in one Hospital | Pituitary adenoma | 1962-1986 | 30 years | 334 | 10-85 years | England and Wales population | Cerebrovascular death from death certificate | Age, sex, calendar period |
| **Olsson et al 2015(2)** | Sweden | Swedish Cancer Registry | Craniopharyngioma | 1997-2011 | 1997-2011, Mean 9 years (0-25 years) | 307 | Childhood onset (35%) & adulthood onset | Swedish general population | Cause of death ICD 10 code including ischemia heart disease and cerebrovascular | Sex, five-year age group, one-year calendar period |
| **Wijnen et al 2018(3)** | Sweden, Netherlands | Hospital based cohort | Craniopharyngioma | 1987-2014 | Median 13 years | Total 224, 144 Dutch, 80 Swedish | Childhood (50%) and adult onset | Swedish general population and Dutch general population | ICD 10 codes mortality and morbidity, cause specific mortality for malignant neoplasm, IHD, cerebrovascular disease) | Sex, five -year age group, one-year calendar period |
| **Yuen et al 2018(4)** | 28 countries | Pfizer International Metabolic Database largest pharmaco-epidemiological surveillance study | Craniopharyngioma | 1994-2012 | Median 5.3 years | 1669(all patients) | Childhood and adult onset | General population from WHO | Medical history for cardiovascular and cerebrovascular mortalities | Age, sex, region |
| **Henson et al 2017(5)** | UK | Teenage and Young Adult cancer survivor study | Multiple cancer site including Brain and CNS including malignant, benign, unspecified tumours | 1971-2006 | Survived at least 5 years from diagnosis | 17280(all cancer patients) | Diagnosed at 15-39 years | England and Wales population | Cardiac death ICD 10 codeI01, I02.0, I05 to I09, I11, I13, I20 to I25, I27.1 to I27.9, and I30 to I52 and ICD 9 codes: 391, 392.0, 393 to 398, 402, 404, 410 to 414, 416, and 420 to 429. | Age, sex, calendar year |

| | | | | | | | | | | |
|---|---|---|---|---|---|---|---|---|---|---|
| **Prasad et al 2012(6)** | Finland | Finnish Cancer Registry | Multiple non-malignant cancer sites including CNS tumours | 1966-1999 | Follow up 1971-2008, survival at least 5 years | 1346(all patients) | Diagnosed before 35 years | Finish population | Death from circulatory disease (ICD I00-I99), ischemic heart disease (codes I20-I25), other disease(codes I30-I52),cerebrovascular diseases and other diseases of the circulatory system. | Age, sex, calenderer year |
| **Zaorsky et al 2019(7)** | US | SEER | Multiple Invasive cancer sites including CNS tumours | 1992-2015 | 20 years cohort follow up | 99434 | All age | US population | Cerebrovascular accident, ICD-9 code 434.11, or ICD-10 code I63.9. | Age, race, sex |

*ICD: International Classification of Disease

**Table 2 Cardiovascular mortality risk in CNS tumours: synthesis of data**

| **Circulatory mortality or cardiac mortality** | | |
|---|---|---|
| **Authors** | **Death (n)** | **SMR(95% CI)*** |
| **Olsson et al 2015(craniopharyngioma)** | 13 | 4.3(2.3-7.4) |
| **Wijnen etal 2018 (craniopharyngioma)** | 8 | 2.3(1.10-4.50) |
| **Yuen et al 2018(craniopharyngioma)** | 18 | 2.0(1.20-3.20) |
| **Henson et al 2016(malignant and non-malignant)** | 144 | 1.4(1.1-1.6) |
| **Prasad et al 2012(malignant)** | 10 | 2.6(1.3-4.8) |
| **Ischemic heart disease mortality** | | |
| **Authors** | **Death (n)** | **SMR(95% CI)** |
| **Olsson et al 2015(craniopharyngioma)** | 7 | 3.7((1.5-7.6) |
| **Wijnen etal 2018 (craniopharyngioma)** | 4 | 1.1(0.4-3.0) |
| **Henson et al 2016(malignant and non-malignant)** | 114 | 1.33(1.1-1.6) |
| **Prasad et al 2012(malignant)** | 1 | 0.60(0.01-3.1) |
| **Cerebrovascular disease or stroke mortality** | | |
| **Authors** | **Death (n)** | **SMR(95% CI)** |
| **Olsson et al 2015(craniopharyngioma)** | 5 | 5.1(1.7-12.0) |
| **Brada et al 2002 (benign)** | 33 | 4.1 |
| **Wijnen etal 2018 (craniopharyngioma)** | 4 | 5.8(2.2-15.5) |
| **Yuen et al 2018(craniopharyngioma)** | 8 | 3.2(1.40-6.30) |
| **Prasad et al 2012(malignant)** | 8 | 8.2(3.5-16.1) |
| **Zaorsky et al 2019(malignant)** | 525 | 7.63(5.66-10.5) |
| ***Standardised mortality ratio (SMR)** | | |

**Summary of results**

We searched OVID MEDLINE and EMBASE for epidemiological studies, reviews, and guidelines published in English from Jan 1998 to Dec 2021, using search terms for brain or central nervous system tumour, and for cardiovascular outcomes. We identified seven articles that calculated risk (either by standardised mortality ratios, SMR, or Hazard ratios, HR) of any specific or composite cardiovascular disease (CVD) mortality outcome among CNS tumour patients. Three studies presented estimates for multiple malignant cancer sites including CNS tumours: four focused on younger age (<40 years) onset tumours. The most common outcomes were stroke/cerebrovascular disease (n=6) and cardiovascular or heart disease(n=4). Elevated CVD mortality risk was observed in most studies, though these studies were limited to younger age onset CNS tumours with small number or reported aggregated data for CNS tumours as an overall group. Limited evidence was reported for non-malignant CNS tumours, particularly meningioma.
